# Phenome-Wide Association of *APOE* Alleles in the *All of Us* Research Program

**DOI:** 10.1101/2024.09.04.24313010

**Authors:** Ehsan Khajouei, Valentina Ghisays, Ignazio S. Piras, Kiana L. Martinez, Marcus Naymik, Preston Ngo, Tam C. Tran, Joshua C. Denny, Travis J. Wheeler, Matthew J. Huentelman, Eric M. Reiman, Jason H. Karnes

## Abstract

**Background:** Genetic variation in *APOE* is associated with altered lipid metabolism, as well as cardiovascular and neurodegenerative disease risk. However, prior studies are largely limited to European ancestry populations and differential risk by sex and ancestry has not been widely evaluated. We utilized a phenome-wide association study (PheWAS) approach to explore *APOE*- associated phenotypes in the *All of Us* Research Program.

**Methods:** We determined *APOE* alleles for 181,880 *All of Us* participants with whole genome sequencing and electronic health record (EHR) data, representing seven gnomAD ancestry groups. We tested association of *APOE* variants, ordered based on Alzheimer’s disease risk hierarchy (ε2/ε2<ε2/ε3<ε3/ε3<ε2/ε4<ε3/ε4<ε4/ε4), with 2,318 EHR-derived phenotypes. Bonferroni-adjusted analyses were performed overall, by ancestry, by sex, and with adjustment for social determinants of health (SDOH).

**Findings:** In the overall cohort, PheWAS identified 17 significant associations, including an increased odds of hyperlipidemia (OR 1.15 [1.14–1.16] per *APOE* genotype group; *P*=1.8×10^-129^), dementia, and Alzheimer’s disease (OR 1.55 [1.40–1.70]; *P*=5×10^-19^), and a reduced odds of fatty liver disease (OR 0.93 [0.90–0.95]; *P*=1.6×10^-9^) and chronic liver disease. ORs were similar after SDOH adjustment and by sex, except for an increased number of cardiovascular associations in males, and decreased odds of noninflammatory disorders of vulva and perineum in females (OR 0.89 [0.84–0.94]; *P*=1.1×10^-5^). Significant heterogeneity was observed for hyperlipidemia and mild cognitive impairment across ancestry. Unique associations by ancestry included transient retinal arterial occlusion in the European ancestry group, and first-degree atrioventricular block in the American Admixed/Latino ancestry group.

**Interpretation:** We replicate extensive phenotypic associations with *APOE* alleles in a large, diverse cohort, despite limitations in accuracy for EHR-derived phenotypes. We provide a comprehensive catalog of *APOE*-associated phenotypes and present evidence of unique phenotypic associations by sex and ancestry, as well as heterogeneity in effect size across ancestry.

**Funding:** Funding is listed in the acknowledgements.

## INTRODUCTION

Apolipoprotein E (*APOE*) is a multifunctional protein with a major role in lipid metabolism and transportation. The three common alleles *APOE* ε3, *APOE* ε4, and *APOE* ε2 are the result of amino acid substitutions in the *APOE* protein at position 112 or 158, caused by two single-nucleotide polymorphisms (SNPs) within exon 4 of the gene (rs429358 and rs7412 respectively)^1–5^. *APOE* variants have been associated with an increased risk of developing certain neurological, cardiovascular, and metabolic conditions such as Alzheimer’s disease (AD), hypertensive and ischemic heart disease, atherosclerosis, stroke, and hyperlipidemia, among others^3,4,6–9^. The ε4 allele is the strongest known common genetic risk factor for developing AD at older ages while ε2 is considered protective with respect to AD and ε3 as having a neutral influence on risk^5,9–15^. Prior studies of *APOE* are predominantly conducted in individuals of European ancestry and rarely include data for social determinants of health (SDOH). This lack of diversity and data limits our understanding of biological versus sociocultural factors underlying disease risk and exacerbates existing health inequalities^16–18^.

Phenome-wide association studies (PheWAS) systematically examine relationships between genetic variation and EHR-derived phenotypes. PheWAS can also be used to examine new evidence of differential risk across demographic groups. Uncovering previously unknown associations and differential phenotypic risks across demographic groups has the potential to improve our understanding of shared mechanisms underlying disease etiology^19–21^. Previously, Lumsden et al. reported a PheWAS of *APOE* allelic variation using UK Biobank data, replicating decades worth of *APOE* associations. However, the UK Biobank contains data predominantly from individuals of European ancestry and Lumsden et al. restricted their study to only White British individuals^20^. The availability of the *All of Us* Research Program (*All of Us*), a large, diverse DNA biobank coupled to electronic health records (EHR) enables investigation of the relationship between genetic variation and many diseases across multiple populations simultaneously^17,22^. *All of Us* also enables investigation of differential phenotypic associations and heterogeneity of effect sizes with *APOE* allelic variation across demographic groups, accompanied by adjustment for SDOH.

## METHODS

### *All of Us* Research Program

*All of Us* has collected physical measurements, survey responses, longitudinal EHR, and genomic data. The curated data repository release version 7 (CDRv7) contains data on 413,450 participants, including EHR data for more than 287,012 participants and short read whole genome sequencing (srWGS) data for 245,388 participants. EHR data originated from more than 50 healthcare providers within the country and was standardized using the Observational Medical Outcomes Partnership (OMOP) Common Data Model^23^. Currently, the program engages adults 18 years old and older who reside in the USA or a US territory. Informed consent for all participants is conducted in person or through an eConsent platform that includes primary consent, HIPAA Authorization for Research use of EHRs and other external health data, and Consent for Return of Genomic Results. The protocol was reviewed by the Institutional Review Board (IRB) of the *All of Us* Research Program. The *All of Us* IRB follows the regulations and guidance of the NIH Office for Human Research Protections for all studies, ensuring that the rights and welfare of research participants are overseen and protected uniformly.

### Study Cohort and Phenotypic Mapping

This study focused on *All of Us* participants with longitudinal EHR data, srWGS data, and inferred genetic ancestry information. Additionally, analysis was restricted to participants that identify as female or male at birth and those with SNP data to determine *APOE* allelic variation (n=245,366). For phenotypic mapping and PheWAS analysis, the PheWAS ToolKit (PheTK) package was utilized in the *All of Us* Researcher Workbench (https://workbench.researchallofus.org)^24^. The PheTK extracts phenotypic data, covariates, and other relevant information for running PheWAS from OMOP compatible databases. Data on age, sex assigned at birth, EHR length (duration of time between first and last EHR index dates), code occurrence count, and condition count were among the retrieved data (Supplementary Figure S1).

International Classification of Disease (ICD) codes (ICD versions 9-CM, 10, and 10-CM) were mapped to phecode version 1.2 and phecodeX 1.0^24^. An individual was considered a case for a particular phecode when assigned two or more instances of a phecode count. Phecodes having fewer than 50 cases or controls were excluded from each analysis, including subgroup analyses. Phecodes were classified in 18 categories including Immunologic, Cardiovascular, Congenital, Dermatological, Metabolic, Gastrointestinal, Genetic, Genitourinary, Infections, Mental, Musculoskeletal, Neonatal, Neoplasms, Neurological, Pregnancy (excluded in the males only analysis), Respiratory, Sense Organs, and Symptoms.

### *APOE* Allelic Variation

We defined *APOE* allelic dose based on combined genotypes for SNPs rs429358 and rs7412^25^. SNP data was retrieved from the *All of Us* Allele Count/Allele Frequency (ACAF) unphased callset, which encompasses variants exceeding a population-specific allele frequency threshold (>1%) or allele count (>100). Genotype extraction was performed using PLINK 1.9^26^. *APOE* genotypes were imputed based on allele combinations using R (R Core Team 2021) (Supplementary Table S1). Variants and allele frequencies for *APOE* were calculated for the overall cohort and for each ancestry and sex sub-group. Out of the 245,394 participants in *All of Us* with *APOE* data, 28 individuals lacked information on one of the determining SNPs, either rs429358 or rs7412, and were excluded from analysis. Additionally, these SNPs were tested for Hardy-Weinberg equilibrium for the whole dataset and for each ancestral groups using PLINK 1.9 (Table S4)^26^.

### Statistical Analysis

Using the PheTK package within the *All of Us* Researcher Workbench, we applied logistic regression to test associations between phecode and *APOE* variants expressed as an ordinal variable using the following hierarchy: ε2/ε2<ε2/ε3<ε3/ε3<ε2/ε4<ε3/ε4<ε4/ε4. In this model, each odds ratio (OR) should be interpreted as the increased odds for a given phecode for each increase in this order of variants. *APOE* allelic variation was an independent variable in PheWAS logistic regression performed iteratively across available phecodes after adjustment for age, sex at birth, and EHR length (Figure S2). To address confounding by population stratification, the first 16 genetic principal components (PCs) available in *All of Us* were added to all association analyses. *All of Us* Research Program Genomics Investigators previously generated these PCs and inferred ancestral groups as previously described^22^. Accordingly, the inferred ancestry groups within *All of Us* are based on gnomAD ancestry group labels, including African/African American (AFR), American Admixed/Latino (AMR), East Asian (EAS), European (EUR), Middle Eastern (MID), Other (OTH), and South Asian (SAS). Briefly, a random forest classifier was trained on a set of 1000 Genomes and Human Genome Diversity Project (HGDP) autosomal samples with ancestral labels. Next, to predict the ancestry, the *All of Us* samples were projected to the classifier. Finally, the resulting PC data were combined with 1000 genomes and HGDP samples to infer individual participant genetic ancestry using Rye^22,23^.

A Bonferroni correction for multiple comparisons based on the number of phenotypes analyzed was used in the overall cohort and each sub-group analyses. For the analyses in the overall cohort a Bonferroni significance level of α=2.16×10^-05^ (*0.05/2,318 phecodes*) was used. Similarly, Bonferroni adjusted cutoffs for the sex sub-group analysis were α=2.9×10^-05^ for males (*0.05/1,727 phecodes*) and 2.39×10^-05^ for females (*0.05/2,096 phecodes*). Analysis breakdown of the seven ancestry sub-groups, including, AMR, EUR, AFR, EAS, MID, OTH, and SAS, were also Bonferroni corrected using the same formula. In further exploratory analyses, we performed PheWAS for increasing copies of ε2 (ε2/ε2<ε2/ε3<ε3/ε3) and ε4 (ε3/ε3<ε3/ε4<ε4/ε4) in the overall cohort. For ease of interpretation of our results relative to prior literature, we also performed analyses comparing *APOE* ε3/ε4 and ε4/ε4 to the reference group ε3/ε3. Furthermore, a sensitivity analysis was performed after adjustment for SDOH factors, including highest education level, household income, and social deprivation index, into the association analysis for the overall cohort (see Supplemental Methods). To assess the heterogeneity of effect sizes by demographic groups, we conducted heterogeneity testing on the 17 top associations from the whole cohort that appeared across subgroup analyses by sex and ancestry group.

## RESULTS

The overall cohort with sufficient genotypic and EHR data consisted of 181,880 participants. On average, EHR length for the cohort spanned 9.68 (standard deviation [SD]=8.34) years (Figure S2). The average age of participants was 56.95 (SD=16.93) years, females were younger with average age of 55.46 (SD=16.84) compared to 59.35 (SD=16.79) years for males (Figure S3). In the overall cohort, the most frequent allele was ε3, with an allele frequency of 0.774 and 109,577 (60.2%) participants characterized as ε3/ε3 (Table 1). The ε4 allele had a frequency of 0.148 and ε2 had a frequency of 0.078. Proportions of *APOE* genotype differed substantially based on ancestry. For instance, the AFR subgroup had the lowest ε3 frequency (0.678) and the highest ε2 (0.107) and ε4 (0.215) frequencies.

**Table 1.** Total number and proportion of *APOE* genotype and allele frequency for the overall cohort, by ancestry group, and by sex assigned at birth.

|  |  | *APOE* genotype frequency |  |  |  |  |  | *APOE* allele frequency |  |  |
| --- | --- | --- | --- | --- | --- | --- | --- | --- | --- | --- |
|  |  | ε2ε2 | ε2ε3 | ε2ε4 | ε3ε3 | ε3ε4 | ε4ε4 | ε2 | ε3 | ε4 |
| **Overall, n (%)** | 181,880 | 1,237 (0.7) | 21,505 (11.8) | 4,373 (2.4) | 109,577 (60.2) | 40,983 (22.5) | 4,205 (2.3) | 7.8% | 77.4% | 14.8% |
| **Ancestry Group<sup>a</sup>** |  |  |  |  |  |  |  |  |  |  |
| AFR | 35,586 (19.6) | 422 (1.2) | 5,157 (14.5) | 1,585 (4.5) | 16,299 (45.8) | 10,501 (29.5) | 1,622 (4.6) | 10.7% | 67.8% | 21.5% |
| AMR | 28,810 (15.8) | 61 (0.2) | 1,959 (6.8) | 279 (1.0) | 20,397 (70.8) | 5,700 (19.8) | 414 (1.4) | 4.1% | 84.1% | 11.8% |
| EAS | 3,247 (1.8) | 25 (0.8) | 428 (13.2) | 61 (1.9) | 2,191 (67.5) | 515 (15.9) | 27 (0.8) | 8.3% | 82.0% | 9.7% |
| EUR | 97,200 (53.4) | 634 (0.7) | 11,973 (12.3) | 2,121 (2.2) | 59,730 (61.5) | 20,894 (21.5) | 1,848 (1.9) | 7.9% | 78.4% | 13.7% |
| MID | 407 (0.2) | n<20<sup>b</sup> (0.2) | 40 (9.8) | n<20<sup>b</sup> (1.0) | 319 (78.4) | 41 (10.1) | n<20<sup>b</sup> (0.5) | 5.7% | 88.3% | 6.0% |
| OTH | 15,038 (8.3) | 89 (0.6) | 1,828 (12.2) | 305 (2.0) | 9,466 (62.9) | 3,065 (20.4) | 285 (1.9) | 7.7% | 79.2% | 13.1% |
| SAS | 1,592 (0.9) | n<20<sup>b</sup> (0.3) | 120 (7.5) | n<20<sup>b</sup> (1.1) | 1,175 (73.8) | 267 (16.8) | n<20<sup>b</sup> (0.4) | 4.6% | 86.0% | 9.4% |
| **Sex At Birth** |  |  |  |  |  |  |  |  |  |  |
| Female | 112,435 (61.8) | 787 (0.7) | 13,229 (11.8) | 2,645 (2.4) | 67,860 (60.4) | 25,375 (22.6) | 2,539 (2.3) | 7.8% | 77.5% | 14.7% |
| Male | 69,445 (38.2) | 450 (0.6) | 8,276 (11.9) | 1,728 (2.5) | 41,717 (60.1) | 15,608 (22.5) | 1,666 (2.4) | 7.9% | 77.3% | 14.9% |

<sup>a</sup>AFR indicates African/African American; AMR, American Admixed/Latino; EAS, East Asian; EUR, European; MID, Middle Eastern; OTH, Other; SAS, South Asian.
<sup>b</sup>The *All of Us* Data and Statistics Dissemination Policy prohibits presenting participant counts of fewer than 20.

Of the 3,492 phecodes extracted for the overall cohort, 2,318 had more than 50 cases in the overall cohort and were included in PheWAS analysis. In the overall cohort, 17 phecodes constituting five disease categories (Endocrine/Metabolic, Neurological, Gastrointestinal, Cardiovascular, Genitourinary) were significantly associated with *APOE* allelic variation after Bonferroni correction in the overall cohort (Table 2, Figures 1-2). *APOE* allelic variation was most strongly associated with hyperlipidemia within the Endocrine/Metabolic category (OR 1.15 [1.14– 1.16] per *APOE* genotype group; *P*=1.8×10^-129^), followed by hypercholesterolemia and pure hypercholesterolemia. The three phecodes associated with the highest ORs were Alzheimer’s disease (AD) (OR 1.55 [1.40–1.70]; *P*=5.0×10^-19^), followed by dementias (OR 1.32 [1.26–1.39]; *P*=7.7×10^-29^), and dementias and cerebral degeneration (OR 1.26 [1.21–1.32]; *P*=1.8×10^-24^). Three conditions showing a significantly reduced risk included chronic liver disease (OR 0.94 [0.93–0.96]; *P*=8.1×10^-9^), fatty liver disease (FLD) (OR 0.93 [0.90–0.95]; *P*=1.6×10^-9^), and noninflammatory disorders of vulva and perineum (OR 0.89 [0.84–0.94]; *P*=1.1×10^-5^). The analysis adjusting for SDOH factors in the whole cohort showed the addition of these features did not significantly impact the results, with this analysis generating the same 17 top hits in a similar order.

**Figure 1.**
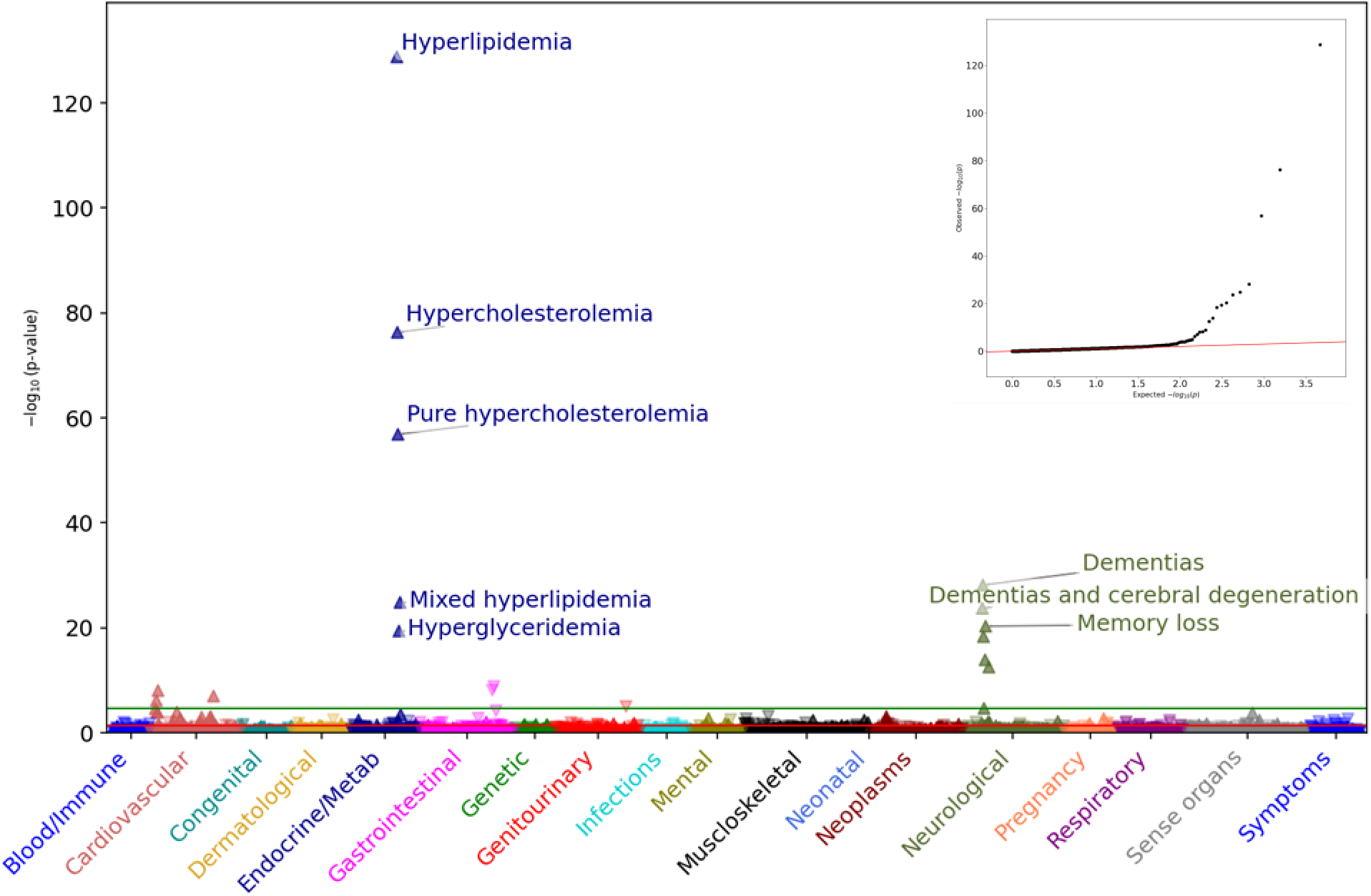
*APOE* PheWAS results from the entire cohort and its accompanying QQ plot. *APOE* genotypes were arranged hierarchically from ε2 to ε4 (ε2/ε2<ε2/ε3<ε3/ε3<ε2/ε4<ε3/ε4<ε4/ε4). PheWAS analysis was adjusted for EHR length, age, sex, and PCs 1-16. In this cohort, 2,318 phecodes with more than 50 cases or controls were analyzed for association with *APOE* variants. The green horizontal line represents Bonferroni correction level (α=2.16×10^-05^) and the red line represents the nominal significance (α=0.05). Furthermore, the direction of arrows indicates either an increased or decreased OR. The x-axis represents the tested phecodes and the broader 18 categories they belong to, and the y-axis shows the -log_10_(*P*-value).

**Figure 2.**
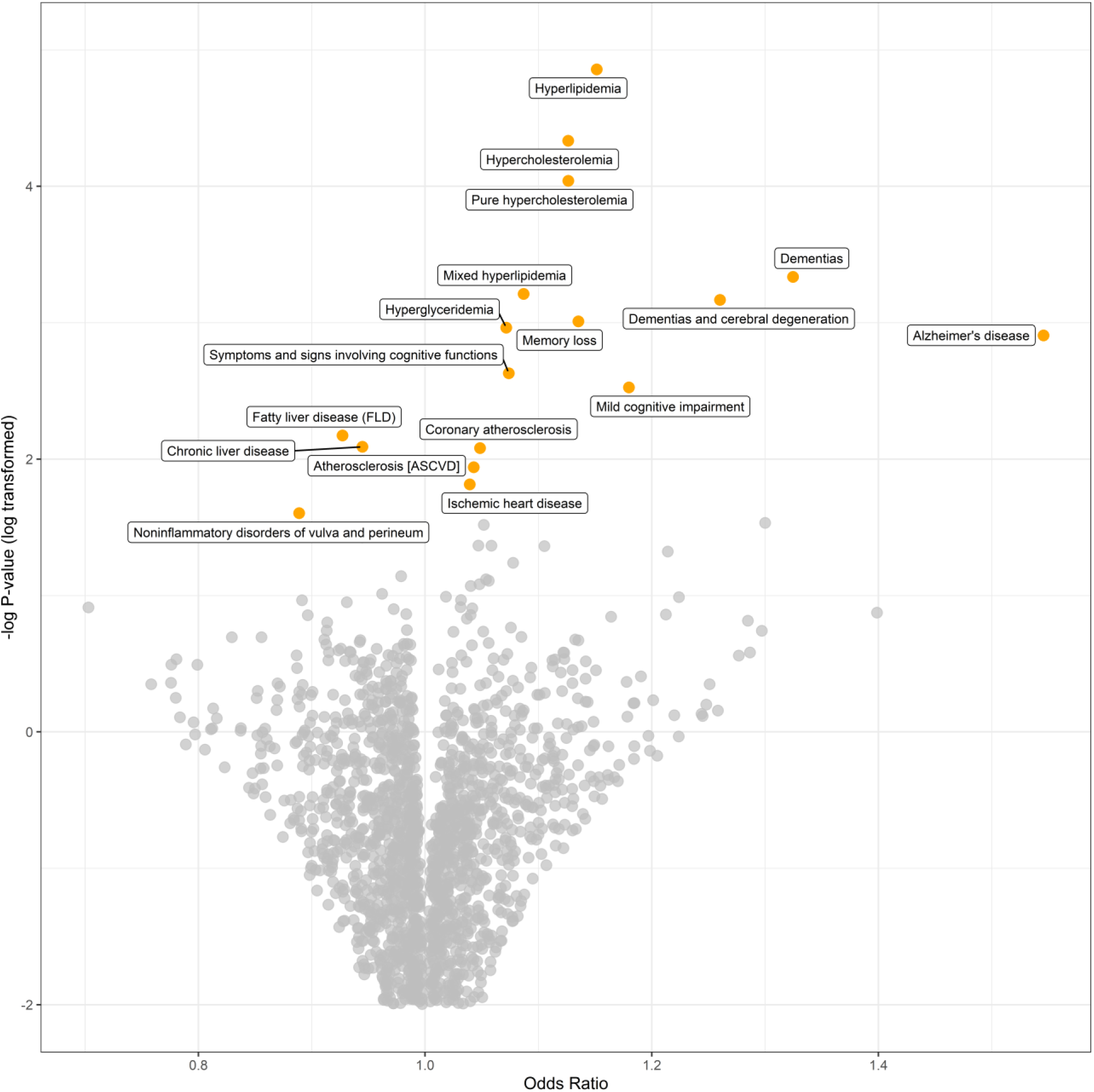
Volcano plot of *P*-values on y-axis against their corresponding odds ratios on x-axis after the PheWAS of the whole cohort. The 17 top hits in this analysis are tagged and labeled in orange. The plot shows AD had the largest effect size while hyperlipidemia had the lowest *P*- value. Moreover, 14 associations are positively associated with *APOE* genotypes, whereas 3 phecodes are inversely associated. N.B. the y-axis is log transformed (*log(-log_10_ (P-value))*) for better visualization with a cut-off at -2.

**Table 2.** Significant associations from PheWAS for the overall cohort (n=181,880).

| Phecode string | Category | Cases/Controls | OR (95% CI) | P |
| --- | --- | --- | --- | --- |
| Hyperlipidemia | Endocrine/Metab | 64,904/105,512 | 1.15 (1.14–1.16) | $1.81 \times 10^{-129}$ |
| Hypercholesterolemia | Endocrine/Metab | 35,178/136,652 | 1.13 (1.11–1.14) | $5.47 \times 10^{-77}$ |
| Pure hypercholesterolemia | Endocrine/Metab | 22,546/150,360 | 1.13 (1.11–1.14) | $1.48 \times 10^{-57}$ |
| Dementias | Neurological | 1,343/179,653 | 1.32 (1.26–1.39) | $7.71 \times 10^{-29}$ |
| Mixed hyperlipidemia | Endocrine/Metab | 17,053/159,320 | 1.09 (1.07–1.10) | $1.53 \times 10^{-25}$ |
| Dementias and cerebral degeneration | Neurological | 1,684/178,586 | 1.26 (1.21–1.32) | $1.77 \times 10^{-24}$ |
| Memory loss | Neurological | 5,139/173,089 | 1.14 (1.11–1.17) | $5.28 \times 10^{-21}$ |
| Hyperglyceridemia | Endocrine/Metab | 19,769/155,567 | 1.07 (1.06–1.09) | $4.35 \times 10^{-20}$ |
| Alzheimer's disease | Neurological | 341/181,310 | 1.55 (1.40–1.70) | $5.02 \times 10^{-19}$ |
| Symptoms and signs involving cognitive functions and awareness | Neurological | 11,576/162,632 | 1.07 (1.05–1.09) | $1.36 \times 10^{-14}$ |
| Mild cognitive impairment (MCI) | Neurological | 1,728/179,148 | 1.18 (1.13–1.23) | $3.24 \times 10^{-13}$ |
| Fatty liver disease (FLD) | Gastrointestinal | 6,770/169,813 | 0.93 (0.90–0.95) | $1.62 \times 10^{-09}$ |
| Chronic liver disease | Gastrointestinal | 10,912/164,358 | 0.94 (0.93–0.96) | $8.14 \times 10^{-09}$ |
| Coronary atherosclerosis [Atherosclerotic heart disease] | Cardiovascular | 17,503/158,757 | 1.05 (1.03–1.07) | $9.64 \times 10^{-09}$ |
| Atherosclerosis [ASCVD] | Cardiovascular | 19,704/155,112 | 1.04 (1.03–1.06) | $1.08 \times 10^{-07}$ |
| Ischemic heart disease | Cardiovascular | 19,903/155,472 | 1.04 (1.02–1.06) | $7.25 \times 10^{-07}$ |
| Noninflammatory disorders of vulva and perineum | Genitourinary | 1,446/108,796 | 0.89 (0.84–0.94) | $1.07 \times 10^{-05}$ |
| Vascular dementia* | Neurological | 214/181,516 | 1.30 (1.15–1.47) | $2.35 \times 10^{-05}$ |
| Angina pectoris* | Cardiovascular | 6,896/170,594 | 1.05 (1.03–1.08) | $2.75 \times 10^{-05}$ |

The PheWAS of the whole cohort was carried out using the PheTK package in the *All of Us* Researcher Workbench. The analysis was adjusted for covariates including age, sex at birth, EHR length data, and 16 genetic PCs. In this logistic regression model, 2,318 phecodes (out of 3,492) met the implemented criteria from which 17 phecodes passed the Bonferroni correction ( $\alpha=2.16 \times 10^{-05}$ ).
\*Nominally significant phenotypic associations that did not pass the significance level set by Bonferroni correction.

In the exploratory analysis for increasing copies of ε2 (ε2/ε2<ε2/ε3<ε3/ε3), all significant associations belonged to the Endocrine/Metabolic category led by hyperlipidemia (OR 1.41 [1.36– 1.46]; *P*=6.6×10^-80^). No signals related to the Neurological category were statistically significant. Moreover, pure hyperglyceridemia was inversely associated in this analysis, suggesting *APOE* ε2 as the risk factor for this condition (Table S2, Figure S6a). In the analysis for increasing copies of ε4 (ε3/ε3<ε3/ε4<ε4/ε4), hyperlipidemia (OR 1.21 [1.18–1.24]; *P*=3.6×10^-49^) was the top association followed by dementia and 13 other phecodes, including significant negative associations with Gastrointestinal phenotypes. In this analysis, Neurological phecodes had larger effect sizes than the overall analysis where all *APOE* variants were included. AD, in particular, showed a relatively high OR (2.47 [2.06–2.97]; *P*=5.9×10^-22^) (Table S3, Figure S6b). We observed similar results in comparisons of ε3/ε3 versus ε3/ε4 (OR 2.07 [1.61–2.65]; *P*=8.8×10^-09^) and ε3/ε3 versus ε4/ε4 (OR 7.42 [4.91–11.21]; *P*=1.8×10^-21^) (Figures S7 and S8).

Female and male sub-group analyses revealed similarities with results from the overall cohort but also some differences that were not exclusively related to sex-specific phenotypes. In females and males, 2,096 and 1,727 phecodes respectively had more than 50 cases or controls and were tested. Similarities included the same top 3 diseases associated with the highest risk in the overall cohort for both sexes (i.e., AD, dementias, and dementias and cerebral degeneration, Figures 3 and S4). However, the top three diseases associated with a reduced risk were only significant in females, including chronic liver disease (OR 0.94 [0.92–0.96]; *P*=9.1×10^-7^), FLD (OR 0.93 [0.90–0.96]; *P*=5.7×10^-6^), and noninflammatory disorders of the vulva and perinium (OR 0.89 [0.84–0.94]; *P*=1.1×10^-5^) (Figure S4a). In contrast, a small but significant increased odds of atherosclerotic heart disease (OR 1.05 [1.03–1.08]; *P*=1.6×10^-6^) and ischemic heart disease (OR 1.05 [1.03–1.07]; *P*=1.2×10^-5^) were observed in males (Figure S4b). Endocrine/Metabolic phecodes, led by hyperlipidemia and hypercholesterolemia, were the strongest associations observed in sub-group analyses for both male and females (Figure S4). Heterogeneity testing indicated heterogenetiy of effect sizes among Endocrine/Metabolic phecodes, including hyperlipidemia, hypercholesterolemia, pure hypercholesterolemia, and mixed hyperlipidemia (Figure 3).

**Figure 3.**
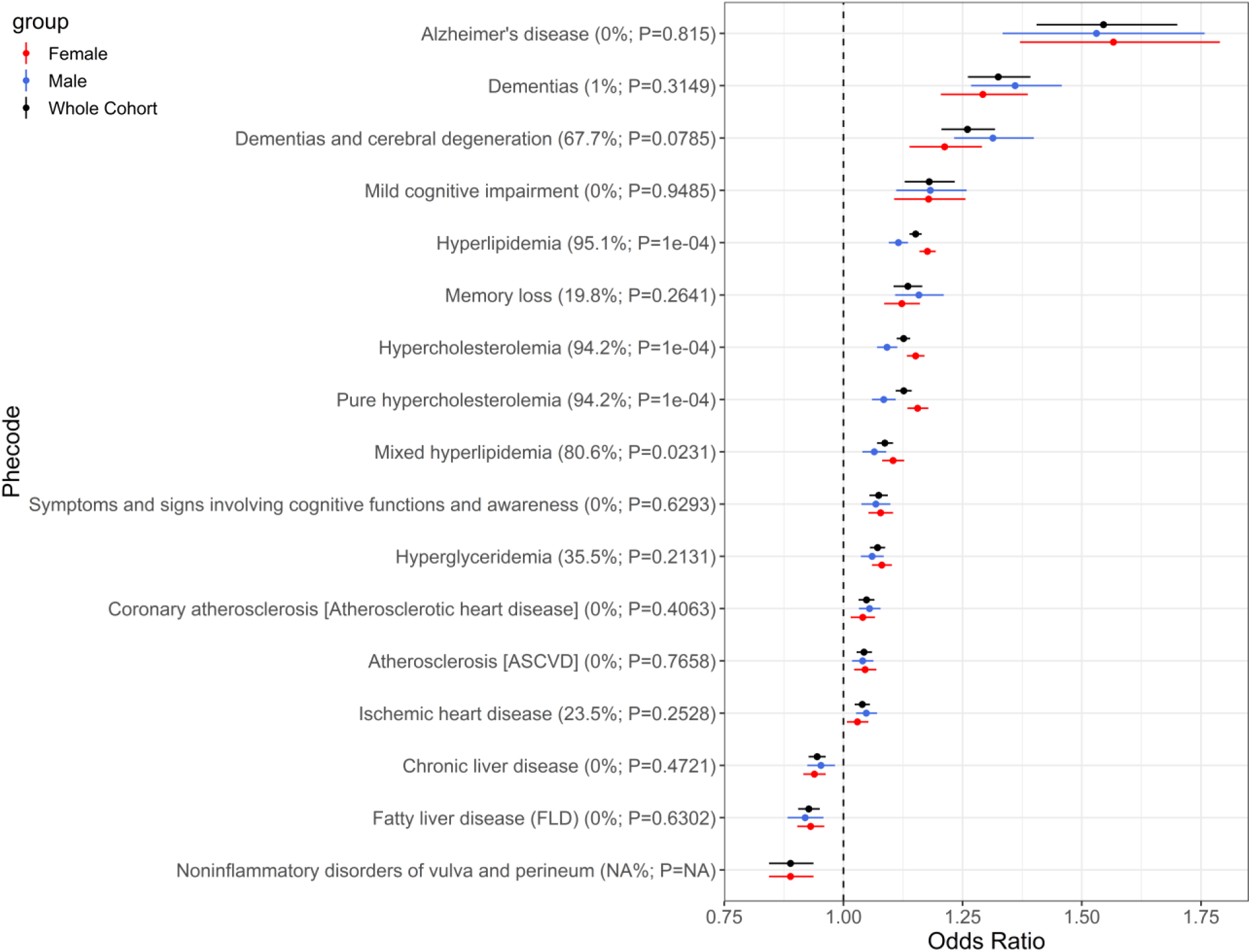
Plot illustrates odds ratios from three separate analyses combined. The top hits from the PheWAS for the whole cohort were used to extract the similar phecodes from PheWAS summary outcomes in females and males. We can see that confidence intervals follow the similar patterns in three analyses and are larger in AD and dementias compared to other phecode categories. The results of heterogeneity test, I^2^ and *P*-value, for each phecode is presented in the label of each corresponding phecode. One phecode, noninflammatory disorders of vulva and perineum, was excluded from the heterogeneity test because it was available for females. Four out of 16 tests indicated heterogeneity in results (*P*-values<0.05). These phecodes were hyperlipidemia, hypercholesterolemia, pure hypercholesterolemia, and mixed hyperlipidemia.

In the ancestral groups AFR, AMR, EAS, EUR, MID, OTH, and SAS, 1,501, 1,398, 430, 2,105, 60, 1,273, and 226 phecodes respectively were tested for association. In the largest group (EUR), the top three diseases associated with the highest risk in descending order were AD (OR 1.67 [1.48–1.89]; *P*=1.0×10^-16^), followed by dementias (OR 1.36 [1.28–1.45]; *P*=4.5×10^-21^), and transient retinal arterial occlusion [Amaurosis fugax] which was unique to the Eur sub group (OR 1.31 [1.16–1.48]; *P*=9.8×10^-6^). Interestingly, trichomoniasis was also exclusively associated within the EUR ancestry group (OR 1.05 [1.03–1.08]; *P*=2.3×10^-5^) (Figures 4 and S5d). However, the regression model related to trichomoniasis did not converge, possibly due to a low number of cases in this phecode (102 cases versus 96,904 controls).

**Figure 4.**
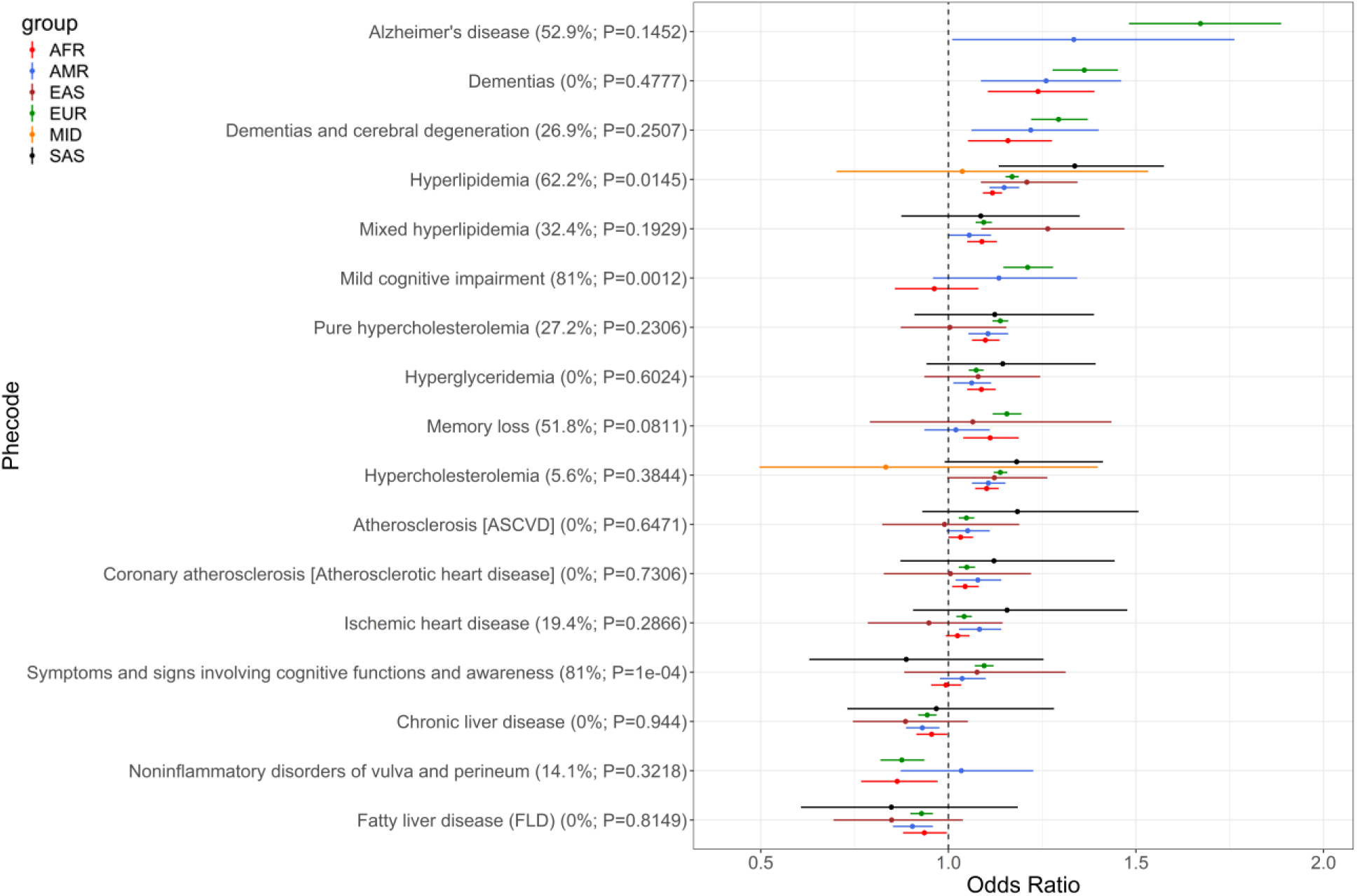
The forest plot displays the combined odds ratios from six separate analyses across ancestral groups. The 17 top hits from the PheWAS for the whole cohort were used to extract the similar phecodes from PheWAS summary outcomes in ancestral groups. Not all the above phecodes had case counts above 50 in all ancestral categories, such as AD appearing only for EUR and AMR. The results of heterogeneity test, I^2^ and *P*-value, for each phecode is presented in the label of the corresponding phecode. Three associations including hyperlipidemia, mild cognitive impairment, and symptoms and signs involving cognitive functions and awareness indicated heterogeneity in results (*P*-values<0.05).

For AMR, only three disease associations survived Bonferroni correction including first-degree atrioventricular block (OR 1.74 [1.38–2.20]; *P*=3.4×10^-6^), followed by hyperlipidemia (OR 1.15 [1.11–1.19]; *P*=1.9×10^-15^), and hypercholesterolemia (OR 1.11 [1.06–1.15]; *P*=9.4×10^-7^) (Figures 4 and S5b). In AFR, only five diseases related to dyslipidemia in the Endocrine/Metabolic category survived multiple corrections including, hyperlipidemia, hypercholesterolemia, pure hypercholesterolemia, mixed hyperlipidemia, and hyperglyceridemia (Figures 4 and S5a). PheWAS for EAS, MID, and SAS did not identify any significant associations, presumably due to the small number of participants in certain *APOE* genotypes, particularly within ε2/2 and ε4/4 (Figure S5c-e-f). We observed significant heterogeneity in 3 out of 17 phecodes tested by ancestry group. Phecodes with heterogenous associations by ancestry included both Endocrine/Metabolic and Neurological categories and consisted of hyperlipidemia, mild cognitive impairment, and symptoms and signs involving cognitive functions and awareness (Figure 4).

## DISCUSSION

We investigated the association between 2,318 clinical diagnostic codes and the six common *APOE* variants from the *All of Us* database. We also performed PheWAS of *APOE* allelic dose by sex at birth and by ancestry and evaluated heterogeneity of associations across these groups. We identified 17 phecodes associated with *APOE* genotypes reflecting Endocrine/Metabolic, Neurological, Gastrointestinal, Cardiovascular, and Genitourinary phecodes (Table 2, Figures 1-4). We observed *APOE* associations with hyperlipidemia and AD, with *APOE* ε4 carrying the highest risk compared to the other variants. Observed associations in European ancestry groups and in females and males largely recapitulated results in the overall cohort. We also observed evidence of unique *APOE* associations by sex and by ancestry, as well as heterogeneity across effects sizes primarily for neurologic phenotypes.

Seminal studies on *APOE* have revealed significant associations between alleles (*APOE* ε2 and *APOE* ε4) and AD and hypercholesterolemia, with notable variation by sex and race/ethnicity. For AD, the *APOE* ε4 allele is linked to increased risk of AD, with ORs ranging from 3-4 for heterozygous individuals and 12-15 for homozygous individuals, with odds significantly increase in neuropathologically confirmed AD cases^3,5,10,28^. In this study the strongest association between AD and *APOE* genotype was accompanied with an OR of 7.42 when only *APOE* ε4 homozygous carriers compared to *APOE* ε3 homozygous from the overall cohort (Figure S8). The reduced OR for AD observed in the present study may be a result of the observational nature of this study, including potential misclassification and/or underdiagnosis of AD. In addition, *All of Us* enrolls only those able to consent and has been open to enrollment only for several years. Therefore, the number of AD cases and ORs associated with *APOE* alleles may increase with the continuation of the program and increased age of participants.

*APOE* variants showed distinct frequencies across different ancestry groups. Participants with MID ancestry had the highest frequency of ε3, whereas those with AFR ancestry had the lowest. On the other hand, ε4 showed an opposite trend, with the highest frequency observed in the AFR group and the lowest in the MID group, consistent with prior results^29^. In addition to Lumsden et al. (2020) who reported PheWAS of *APOE* alleles using UK Biobank data, Belloy et al. (2023) examined the association between *APOE* genotype and AD across age, sex, and both race/ethnic and ancestral groups including European, African, and Amerindian. In both studies a genetic model, where the association of a phecode and each *APOE* variant was estimated with reference to *APOE* ε3ε3, was applied. These publications reported ORs of 13.5 and 12.84, respectively, for the association between *APOE* ε4ε4 and AD in individuals of European ancestry. Similarly, the reported effect sizes were 3.69 and 3.43 in case of *APOE* ε3ε4 association. Effect size estimates were reduced for African and Amerindian individuals^20,28^.

In the present publication, we observed similarly attenuated ORs in non-European ancestry groups that had sufficient case counts for analysis. In contrast to prior publications, our primary analysis applied an additive model where all six *APOE* variants were simultaneously included in each association analysis with a phecode. Moreover, we were able to perform an *APOE* allelic dose PheWAS across seven available ancestry groups as well as across sex. Lumsden et al. (2020) reported OR estimates 1.35 and 1.17 for hypercholesterolemia for *APOE* ε4ε4 and ε3ε4, respectively. Hypercholesterolemia showed an OR of 1.14 in the EUR population of the present study. Our results for the AMR ancestries diverged from those in the whole cohort and the EUR sub-group. Cardiovascular diseases were also positively associated with *APOE* variants, including atrioventricular (AV) block, followed by hyperlipidemia, and hypercholesterolemia. To our knowledge, no prior research has uncovered an association between *APOE* and AV block. This association can be considered hypothesis generating and was not observed in any other ancestry group. This AV block association may be related to increased cardiovascular risk or donepezil usage in AD patients^30^. Since this study examined differential risk across ancestry groups, we evaluated the impact of adjustment for SDOH factors on *APOE* association. Although we did not evaluate the direct effect of SDOH factors on disease risk, our results suggest that SDOH factors had little effect on *APOE*-associated disease risk.

Women with *APOE* ε4 generally have higher ORs compared to men. However, this risk varies across studies, suggesting an age-by-sex interaction^3,10,31–33^. Our study did not confirm a sex specific effect in association between *APOE* genotype and AD, as ORs from female and male subgroup analyses were similar and no evidence of heterogeneity across sex was observed. Sex-specific analyses revealed similarities with the overall cohort but also some differences not exclusively related to sex-specific diagnoses. The three diseases associated with a reduced risk were only seen in females, including chronic liver disease, FLD, and noninflammatory disorders of the vulva and perinium. In contrast, males had significant associations with atherosclerotic heart disease and ischemic heart disease. Lumsden et al. (2020) and others have reported a protective effect of ε4 against chronic liver disease and cirrhosis using the UK Biobank data^20,34^. Furthermore, although not specifically related to noninflammatory disorders of the vulva and perinium, there is evidence in the literature showing associations linking cervical disorders and *APOE*^20^.

There were several limitations to our study. There were reduced sample sizes in the EAS, MID, and SAS ancestry groups, compromising statistical power for phenotypes analyzed and reducing the number of phenotypes tested. This was also true to a lesser extent for AFR, OTH, and AMR ancestry groups. Our utilization of EHR data may have resulted in reduced diagnostic accuracy, especially when characterizing cases and controls for neurodegenerative disease such as AD. This may have led to underestimated ORs particularly in AD due to misdiagnosed cases, inclusion of controls who may end up having preclinical AD, and in diagnosis of AD vs non-AD pathology (e.g. cerebrovascular disease). Scalable blood-based biomarker endophenotypes may help to address this challenge and more accurately characterize the presence or absence of AD^5,35^. Similarly, site-specific patterns in billing practices, data upload, and practice patterns, including potential differences in cognitive and clinical assessments, might have confounded results. Although we adjusted for several major SDOH factors, our investigation of the SDOH influences on *APOE*-related disease risk was not comprehensive and limited to those factors that were assessed across the majority of *All of Us* participants. Inclusion of more detailed SDOH data in future studies would aid in disentangling biological versus sociocultural influences on disease. Finally, we did not evaluate carriage of the extremely rare *APOE* ε1 allele. Even in our large cohort, we would not expect to have power to detect *APOE* ε1 associations, since a prior study in 2020 has reported 15 and 2 individuals with respectively ε1ε4 and ε1ε2 genotypes (out of 488,377 individuals) and according to a review study in 2019 only four individuals with *APOE* ε1 genotype have been reported in the literature^4,20^.

In summary, we replicate extensive phenotypic associations with *APOE* alleles in a large, diverse cohort. We provide a comprehensive catalog of *APOE*-associated phenotypes and present evidence of unique phenotypic associations by sex and by ancestry, as well as heterogeneity in effect size across ancestry groups. We confirm recent literature suggesting a reduced risk of FLD with increasing *APOE* ε4 allelic dose, and we identify multiple hypothesis-generating *APOE* allelic dose associations, such as noninflammatory disorders of the vulva and perinium in females and AV block in the AMR ancestry group. Replication of these hypothesis-generating results is needed, as is increased sample sizes for the smaller ancestry groups.

## Supporting information

Supplemental PheWAS Results

## Data Availability

All data and source code for analysis for this project is available within the *All of Us* Researcher Workbench and will be made available to any approved *All of Us* researcher.

https://workbench.researchallofus.org/workspaces/aou-rw-134b018e/apoephewas/

## Acknowledgements

We gratefully acknowledge *All of Us* participants for their contributions, without whom this research would not have been possible. We also thank the National Institutes of Health’s *All of Us* Research Program for making available the participant data examined in this study. The *All of Us* Research Program is supported by the National Institutes of Health, Office of the Director: Regional Medical Centers: 1 OT2 OD026549; 1 OT2 OD026554; 1 OT2 OD026557; 1 OT2 OD026556; 1 OT2 OD026550; 1 OT2 OD 026552; 1 OT2 OD026553; 1 OT2 OD026548; 1 OT2 OD026551; 1 OT2 OD026555; IAA #: AOD 16037; Federally Qualified Health Centers: HHSN 263201600085U; Data and Research Center: 5 U2C OD023196; Biobank: 1 U24 OD023121; The Participant Center: U24 OD023176; Participant Technology Systems Center: 1 U24 OD023163; Communications and Engagement: 3 OT2 OD023205; 3 OT2 OD023206; Community Partners: 1 OT2 OD025277; 3 OT2 OD025315; 1 OT2 OD025337; 1 OT2 OD025276; and the Driver Project Subaward: 1 OT2 OD036485-01. JHK is funded by the NIH’s Office of the Director under awards 1OT2OD026549 and OT2OD036485 and the National Heart, Lung, and Blood Institute (NHLBI) under awards R21HL172036, R01 HL156993, and R01 HL158686. TCT and JCD are supported by the National Human Genome Research Institute (NHGRI) grant HG200417. The authors would also like to thank the National Institute of Health and Arizona ADRC: P30AG072980, the National Institute of Aging: R01AG069453, the State of Arizona AARC: CTR057001 and Arizona Alzheimer’s Consortium.

## Author Contributions

J.H.K., M.J.H. and E.M.R conceived the study; V.G., P.N., and E.K. performed literature review; I.S.P and M.N. extracted the *APOE* genotypic data in *All of Us*; E.K. and K.L.M. performed the PheWAS in *All of Us*; the PheWAS pipeline was created by K.L.M in collaboration with T.C.T., J.C.D., and T.W.; E.K. performed the data analysis and generated the tables and graphs; V.G. and E.K. wrote the manuscript with substantial input from J.H.K, E.M.R, and all authors.

## Availability of data and materials

The data use agreement in *All of Us* prohibits researchers from sharing row-level data in any form. Data from *All of Us* is accessible only through the *All of Us* Researcher Workbench as stipulated in the informed consent of participants in the program. All data and source code for analysis for this project is available within the *All of Us* Researcher Workbench and will be made available to any approved *All of Us* researcher (https://workbench.researchallofus.org/workspaces/aou-rw-134b018e/apoephewas/). Detailed descriptions of cohorts, *APOE* allelic dose analysis, genomic quality control, statistical analyses, and results not presented in the main text are available in the Supplemental Materials.

