## Supplementary Materials for "Phenome-Wide Association of *APOE* Alleles in the *All of Us* Research Program"

**Short title:** *APOE* PheWAS in *All of Us*

Ehsan Khajouei[1*], Valentina Ghisays[2*], Ignazio S. Piras[3], Kiana L. Martinez[1], Marcus Naymik[3], Preston Ngo[1], Tam C. Tran[4], Joshua C. Denny[4,5], Travis J. Wheeler[1], Matthew J. Huentelman[3], Eric M. Reiman[2*], Jason H. Karnes[1,6*]
*Contributed equally

**Affiliations:**
1 Department of Pharmacy Practice and Science, R. Ken Coit College of Pharmacy, University of Arizona, Tucson, AZ, USA
2 Banner Alzheimer's Institute, Phoenix, AZ, USA
3 Neurogenomics Division, Translational Genomics Research Institute, Phoenix, AZ, USA
4 National Human Genome Research Institute, National Institutes of Health, Bethesda, MD, USA
5 All of Us Research Program, National Institutes of Health, Bethesda, MD, USA
6 Department of Biomedical Informatics, Vanderbilt University Medical Center, Nashville, TN, USA

**Corresponding Author:**
Jason H. Karnes, PharmD, PhD
Associate Professor
University of Arizona R. Ken Coit College of Pharmacy
1295 N Martin AVE
Tucson, AZ 85721
520-626-1447


### SUPPLEMENTARY METHODS

*Study Cohort*

The *All of Us* participants with longitudinal EHR data, short read whole genome sequencing (srWGS) data, and inferred genetic ancestry information were included in this study. Figure S1 illustrates a step wise approach of creating the study cohort within the *All of Us* Researcher Workbench. First, a table linking individual IDs to their *APOE* gene variants was generated. This table included genotypic information for 245,366 participants out of the total 245,394. Therefore, there were 28 individuals whose *APOE* genotypes could not be distinguished with the available sequencing and genotypic information. Some of these individuals might belong to the extremely rare *APOE* genotypes, *APOE* ε1 allele which has the combination of an arginine at position 112 and a cysteine at position 158. A PheWAS study on *APOE* data from the UK Biobank reported 15 and 2 individuals with respectively ε1ε4 and ε1ε2 genotypes and a review study reported 4 individuals carrying *APOE* ε1 across the literature[1,2].

The next step involved building the cohort and extracting covariates. A Python-based package called PheWAS ToolKit (PheTK) was used for this purpose. The package, moreover, can run the phenome-wise association analysis. To build a cohort, PheTK requires a list of participant IDs as input. The package extracted information about age, sex assigned at birth, EHR length, code occurrence count, and condition count on 181,880 participants from several tables. This is the number of participants in *All of Us* who jointly had the *APOE* and EHR data and recognized themselves as either female or male at birth.

There are more than two possible biological sex categories assigned at birth in *All of Us*. PheTK only included participants identified as female or male, excluding categories such as intersex, or “prefer not to answer”. The extracted EHR data reflects the duration of time between first and last EHR index dates. The code occurrence count refers to the number of times a specific medical code was assigned to a participant. The condition count, on the other hand, indicates the total number of distinct conditions an individual was diagnosed with over the time. Additionally, PheTK extracted the estimated genetic ancestry information alongside the first 16 genetic principal components (PCs) available in *All of Us*.

*All of Us* provides other demographic information on 413,457 individuals about race, gender, and ethnicity. PheTK did not extract this information. Therefore, as a final step, we retrieved this information using SQL query and added it to the table that PheTK already prepared.

*R script used in APOE imputation*

Below is the R script which was utilized to impute the *APOE* variants:

```
# importing package
library(stringr)

# reading table with genotypes
apoe.status.df = read.csv('apoe_status_table.csv',stringsAsFactors = F) # this refers to Table S1
apoe.status.df$full = paste0(apoe.status.df$rs429358,apoe.status.df$rs7412)

# reading plink ped file with genotypes
apoe.df = read.table('apoe.ped', sep=' ',stringsAsFactors = F)
apoe.df$V7V8 <- paste0(apoe.df$V7, apoe.df$V8)
apoe.df$V9V10 <- paste0(apoe.df$V9, apoe.df$V10)

apoe.df <- dplyr::select(apoe.df, V2, V7V8, V9V10)

names(apoe.df)[c(2,3)]=c('rs429358','rs7412')
```

```
apoe.df[apoe.df == 'TC'] ='CT'

# apoe determination
apoe.df$apoe.status = 'NA'
n=0

for (ind in 1:nrow(apoe.df)) {
print(ind)
  snp1 = apoe.df[ind,'rs429358']
  snp2 = apoe.df[ind,'rs7412']
  snps.both = paste0(snp1,snp2)

  apoe.e.status =  apoe.status.df$status[match(snps.both,apoe.status.df$full)]
  apoe.df[ind,'apoe.status'] = apoe.e.status

  n=n+1
}

names(apoe.df)[1]='sample'
print("writing final file")
write.csv(apoe.df,'APOE_status.csv',quote = F,row.names=F)
```

*Statistical analysis*

In this study we examined associations between extracted phecodes and the *APOE* gene variants. The *APOE* variants were treated as an ordinal variable reflecting their previously published Alzheimer's disease risk hierarchy. This ranking increased incrementally from ε2/ε2 towards ε4/ε4 in this fashion: ε2/ε2<ε2/ε3<ε3/ε3<ε2/ε4<ε3/ε4<ε4/ε4. In the analysis, each variant was numbered according to its rank in the combination. The specific numbers assigned to each ranking did not affect the association results, highlighting that the relative order of the rankings was more important than the actual numbers used. For instance, the PheTK produced similar results when we tried different numbering approaches, such as 0, 1, 2, 3, 4, and 5 versus 3, 4, 5, 6, 7, and 8. Furthermore, to explore the effect of available covariates in the association, several trial analyses were conducted in the Researcher Workbench. Among the tested covariates, the code occurrence count, and the condition count slightly affected the association outcome. However, the covariate combination that efficiently controlled false discoveries were age, sex assigned at birth, EHR length, and the first 16 genetics PCs. Therefore, to keep the analysis statistically parsimonious, other covariates were excluded from the model.

*Additional association analyses for ε2 and ε4*

To decipher the outcomes of the above analysis using all the six *APOE* variants, separate analyses were conducted using smaller subsets of the *APOE* variants. First, an incremental increase in the *APOE* gene dosage from ε2 towards ε3 was considered, i.e. ε2/ε2<ε2/ε3<ε3/ε3. Next, we included an increase in gene dosage from ε3 to ε4, considered in this way: ε3/ε3<ε3/ε4<ε4/ε4. Another plausible ranking was to include ε2/ε2, ε2/ε4, and ε4/ε4. However, this combination had the smallest number of participants. To ensure statistically reliable results with sufficient power, we excluded this subgroup from further analysis.

Similar to the main PheWAS using the six *APOE* variants, the PheTK package within the Researcher Workbench was used to test association between the forgoing subgroups of the *APOE* variants and the extracted phecodes in the whole cohort. Age, sex at birth, and EHR length were included in these logistic regression analyses as covariates. Additionally, the first 16 genetic principal components (PCs) available in *All of Us* were added to the analyses to address confounding by population stratification. Furthermore, a Bonferroni correction for multiple testing based on the number of tested phecodes was used in these additional analyses. The correction level was $\alpha=2.27\times10^{-05}$ (*0.05/2,202 phecodes*) when gene dosage from ε2 towards ε3 was considered (ε2/ε2<ε2/ε3<ε3/ε3). Likewise, we applied a significance level of $\alpha=2.2\times10^{-05}$ (*0.05/2,269 phecodes*) to the ε3 and ε4 analysis (ε3/ε3<ε3/ε4<ε4/ε4).

*Complementary analyses for ε3ε4 and ε4ε4 with ε3ε3 as the reference*

In the first analysis, participants from the whole cohort with either *APOE* ε3ε4 and ε3ε3 variants were included. Here (ε3/ε3<ε3/ε4) *APOE* ε3ε4 was assigned a rank of 1 while ε3ε3 was considered 0. For a total number of 150,560 participants, 3,479 phecodes were extracted. The analysis was carried out on 2,251 phecodes with more than 50 cases or controls adjusting for age, sex assigned at birth, EHR length, and the first 16 genetics PCs. A Bonferroni significance level of $\alpha=2.22\times10^{-05}$ (*0.05/2,251 phecodes*) was applied for multiple testing.

In the second analysis 113,782 participants carrying either *APOE* ε4ε4 or ε3ε3 (ε3/ε3<ε4/ε4), were included. For this group, 3,453 phecodes were extracted, of which 2,141 phecodes met the inclusion criteria and were tested for association with *APOE* variants. The included covariates were like the previous analysis with *APOE* ε3ε3 as the reference was ranked 0 and ε4ε4 was ranked 1. Furthermore, a Bonferroni correction level of $\alpha=2.34\times10^{-05}$ (*0.05/2,141 phecodes*) determined the significant associations.

*Analysis adjusting for Social Determinants of Health*

The *All of Us* Research Program provides Social Determinants of Health (SDOH) data for 413,174 participants within a table called “zip code socioeconomic”. This table contains information, among other features, on zip code, high school education, income, and a deprivation index. According to the *All of Us* v7 Data Dictionary, the deprivation index is the population-weighted average of the index for the census tracts covered by the 3-digits ZIP Code Tabulation Areas which are approximate area representations of the US Postal Service ZIP Code. Merging the zip code socioeconomic table with the cohort dataset resulted in a dataset of 181,765 individuals, indicating a lack of SDOH information for 115 participants in the whole cohort. The association analysis was carried out as previously done for the whole cohort carrying the six *APOE* variants. In addition to the previous covariates, we added three SDOH factors including highest education level, household income, and social deprivation index to the model as covariates. A total number of 3,491 phecodes were extracted for the 181,765 participants, out of which 2,319 phecodes had more than 50 cases or controls and were tested for association with the *APOE* variants. Additionally, a Bonferroni correction level of $\alpha=2.16\times10^{-05}$ (*0.05/2,319 phecodes*) was applied to determine the significance level of associations.

*Hardy-Weinberg Equilibrium*

Two single nucleotide polymorphisms (SNPs) which determine the *APOE* variants were investigated for Hardy-Weinberg equilibrium. The SNP information can be found in a callset called ACAF (stands for Allele Count/Allele Frequency threshold) within the Researcher Workbench. First, we extracted SNPs information (rs429358: 19-44908684-T-C and rs7412: 19-44908822-C-T) using a Python-based package, called Hail, from ACAF callset. This callset contains SNPs data for 245,394 participants. Following extraction, the data was exported to PLINK binary data format for analysis[3]. We analyzed SNPs for HWE using PLINK 1.9 for the whole cohort (181,880 participants) of this study in addition to each ancestral group. Table S4 presents the results of this analysis.

*Heterogeneity testing*

We conducted association analysis on the entire cohort and then on several subsets based on sex at birth and the ancestral groups. To evaluate the consistency of our association findings across subgroups (sex and ancestry) compared to the entire cohort, we performed heterogeneity test. The meta package in R was utilized to investigate the existence of heterogeneity in our results[4]. One of the functions within this package requires OR and the corresponding confidence intervals (CIs) form subgroup analyses as input to perform the test. It then performs meta-analysis on the provided test results and produces overall OR and CIs in addition to heterogeneity test results. The heterogeneity test was performed on each phecode that appeared on the 17 top associations from the whole cohort. This was done separately for subgroup analyses for sex (two groups: female and male) and next for ancestral groups (seven groups including African/African American, American Admixed/Latino, East Asian, European, Middle Eastern, Other, and South Asian). First, the PheWAS results for a certain phecode were extracted from subgroup analyses and provided as input to the function in the meta package. Subsequently, the heterogeneity results, $I^2$ and corresponding *P*-values, were tabulated for the top associated phecodes separately for sex and ancestral groups. These results are presented in Figures 3 and 4, respectively for the sex and ancestral groups.

### Supplementary Tables

**Table S1.** Common *APOE* variants and corresponding SNP alleles.

| *APOE* Genotype | rs429358 | rs7412 |
|---|---|---|
| ε1ε1 | CC | TT |
| ε1ε2 | CT | TT |
| ε2ε4 | CT | CT |
| ε1ε4 | CC | CT |
| ε2ε2 | TT | TT |
| ε2ε3 | TT | CT |
| ε3ε3 | TT | CC |
| ε3ε4 | CT | CC |
| ε4ε4 | CC | CC |

The genotypic data source was SNP data called ACAF (Allele Count/Allele Frequency) derived from srWGS data provided on 245,394 individuals in *All of Us*. Following genotype extraction using PLINK, the *APOE* variants were imputed using R script which can be found in the Supplementary Methods[3-5].

**Table S2**. Association results of ε2/ε2<ε2/ε3<ε3/ε3 *APOE* variants in the whole cohort after Bonferroni correction. Among the 2,202 tested phecodes, five of them met the Bonferroni correction level in this analysis.

| Phecode string | Category | Cases/Controls | OR (95% CI) | *P* |
|---|---|---|---|---|
| Hyperlipidemia | Endocrine/Metab | 46,648/77,358 | 1.41 (1.36–1.46) | $6.59 \times 10^{-80}$ |
| Hypercholesterolemia | Endocrine/Metab | 25,250/99,806 | 1.33 (1.28–1.39) | $1.70 \times 10^{-44}$ |
| Pure hypercholesterolemia | Endocrine/Metab | 16,140/109,743 | 1.34 (1.28–1.41) | $4.07 \times 10^{-34}$ |
| Pure hyperglyceridemia | Endocrine/Metab | 2,558/128,023 | 0.64 (0.58–0.69) | $1.08 \times 10^{-25}$ |
| Mixed hyperlipidemia | Endocrine/Metab | 12,270/116,058 | 1.20 (1.14–1.26) | $2.91 \times 10^{-12}$ |
| Occlusion and stenosis of precerebral arteries* | Cardiovascular | 2,601/127,674 | 1.23 (1.11–1.36) | $9.57 \times 10^{-05}$ |
| Ischemic heart disease* | Cardiovascular | 14,483/113,058 | 1.09 (1.04–1.14) | $4.90 \times 10^{-04}$ |

The PheWAS of ε2/ε2<ε2/ε3<ε3/ε3 *APOE* variants in the whole cohort was carried out using the PheTK package in the *All of Us* Researcher Workbench. The analysis was adjusted for covariates including age, sex at birth, EHR length data, and 16 genetic PCs. The number of included participants was 132,319 in the whole cohort for this subset of analysis. Furthermore, 3,468 phecodes were extracted for the whole cohort out of which 2,202 with more than 50 cases or controls met the inclusion criteria.

*Phecodes did not meet the significance level set by Bonferroni correction $\alpha=2.27\times10^{-05}$ (*0.05/2,202 phecodes*).

**Table S3.** Association results of ε3/ε3<ε3/ε4<ε4/ε4 *APOE* variants in the whole cohort after Bonferroni correction. Among the 2,269 tested phecodes, 15 of them met the Bonferroni correction level in this analysis.

| Phecode string | Category | Cases/Controls | OR (95% CI) | *P* |
|---|---|---|---|---|
| Hyperlipidemia | Endocrine/Metab | 56,475/88,488 | 1.21 (1.18–1.24) | $3.60 \times 10^{-49}$ |
| Dementias | Neurological | 1,176/152,819 | 1.81 (1.64–2.00) | $3.66 \times 10^{-31}$ |
| Hypercholesterolemia | Endocrine/Metab | 30,747/115,333 | 1.18 (1.14–1.21) | $4.90 \times 10^{-31}$ |
| Dementias and cerebral degeneration | Neurological | 1,460/151,913 | 1.66 (1.51–1.81) | $3.74 \times 10^{-27}$ |
| Pure hypercholesterolemia | Endocrine/Metab | 19,768/127,218 | 1.17 (1.13–1.21) | $2.90 \times 10^{-22}$ |
| Alzheimer's disease | Neurological | 306/154,255 | 2.47 (2.06–2.97) | $5.88 \times 10^{-22}$ |
| Memory loss | Neurological | 4,466/147,171 | 1.30 (1.23–1.37) | $1.31 \times 10^{-19}$ |
| Symptoms and signs involving cognitive functions and awareness | Neurological | 9,921/138,302 | 1.17 (1.13–1.22) | $1.33 \times 10^{-15}$ |
| Hyperglyceridemia | Endocrine/Metab | 17,071/132,086 | 1.14 (1.10–1.18) | $2.02 \times 10^{-15}$ |
| Mild cognitive impairment | Neurological | 1,505/152,397 | 1.44 (1.31–1.58) | $3.30 \times 10^{-14}$ |
| Mixed hyperlipidemia | Endocrine/Metab | 14,880/135,110 | 1.13 (1.09–1.17) | $3.16 \times 10^{-12}$ |
| Chronic liver disease | Gastrointestinal | 9,316/139,794 | 0.88 (0.84–0.92) | $9.86 \times 10^{-09}$ |
| Fatty liver disease (FLD) | Gastrointestinal | 5,773/144,448 | 0.86 (0.82–0.91) | $1.72 \times 10^{-07}$ |
| Vascular dementia | Neurological | 182/154,453 | 1.85 (1.45–2.37) | $8.70 \times 10^{-07}$ |
| Pure hyperglyceridemia | Endocrine/Metab | 2,840/149,962 | 1.19 (1.11–1.28) | $1.60 \times 10^{-06}$ |
| Chronic nonalcoholic liver disease* | Gastrointestinal | 4,231/147,960 | 0.87 (0.82–0.93) | $2.46 \times 10^{-05}$ |
| Coronary atherosclerosis [Atherosclerotic heart disease]* | Cardiovascular | 14,975/135,002 | 1.08 (1.04–1.12) | $5.57 \times 10^{-05}$ |
| Noninflammatory disorders of vulva and perineum* | Genitourinary | 1,192/92,697 | 0.78 (0.69–0.89) | $1.01 \times 10^{-04}$ |

The PheWAS of ε3/ε3<ε3/ε4<ε4/ε4 *APOE* variants in the whole cohort was carried out using the PheTK package in the *All of Us* Researcher Workbench. The analysis was adjusted for covariates including age, sex at birth, EHR length data, and 16 genetic PCs. The number of included participants was 154,765 in the whole cohort for this subset of analysis. Furthermore, 3,480 phecodes were extracted for the whole cohort out of which 2,269 with more than 50 cases or controls met the inclusion criteria.

*Phecodes did not meet the significance level set by Bonferroni correction $\alpha=2.2\times10^{-05}$ (*0.05/2,269 phecodes*).

**Table S4.** Results from Hardy-Weinberg equilibrium testing performed in each ancestral group and the whole cohort using PLINK 1.9[3].

| Data Subset | SNP | Geno | O(Het) | E(Het) | *P* |
|---|---|---|---|---|---|
| African/African American (AFR) | chr19:44908684:T:C[a] | 1,622/12,086/21,878 | 0.3396 | 0.338 | $3.71 \times 10^{-01}$ |
| | chr19:44908822:C:T[b] | 422/6,742/28,422 | 0.1895 | 0.1905 | $3.30 \times 10^{-01}$ |
| American Admixed/Latino (AMR) | chr19:44908684:T:C | 414/5,979/22,417 | 0.2075 | 0.2084 | $4.97 \times 10^{-01}$ |
| | chr19:44908822:C:T | 61/2,238/26,511 | 0.07768 | 0.07856 | $6.06 \times 10^{-02}$ |
| East Asian (EAS) | chr19:44908684:T:C | 27/576/2,644 | 0.1774 | 0.1752 | $5.48 \times 10^{-01}$ |
| | chr19:44908822:C:T | 25/489/2,733 | 0.1506 | 0.1522 | $5.63 \times 10^{-01}$ |
| European (EUR) | chr19:44908684:T:C | 1,848/23,015/72,337 | 0.2368 | 0.237 | $7.25 \times 10^{-01}$ |
| | chr19:44908822:C:T | 634/14,094/82,472 | 0.145 | 0.1456 | $2.34 \times 10^{-01}$ |
| Middle Eastern (MID) | chr19:44908684:T:C | n<20[c]/45/360 | 0.1106 | 0.1131 | $6.48 \times 10^{-01}$ |
| | chr19:44908822:C:T | n<20/44/362 | 0.1081 | 0.1066 | $1.00 \times 10^{+00}$ |
| Other (OTH) | chr19:44908684:T:C | 285/3,370/11,383 | 0.2241 | 0.2277 | $5.76 \times 10^{-02}$ |
| | chr19:44908822:C:T | 89/2,133/12,816 | 0.1418 | 0.1419 | $9.54 \times 10^{-01}$ |
| South Asian (SAS) | chr19:44908684:T:C | n<20/285/1,300 | 0.179 | 0.1702 | $3.84 \times 10^{-02}$ |
| | chr19:44908822:C:T | n<20/138/1,449 | 0.08668 | 0.08864 | $3.86 \times 10^{-01}$ |
| Overall | chr19:44908684:T:C | 4,205/45,356/132,319 | 0.2494 | 0.2519 | $1.85 \times 10^{-05}$ |
| | chr19:44908822:C:T | 1,237/25,878/154,765 | 0.1423 | 0.1437 | $2.21 \times 10^{-05}$ |

[a]The major allele is T and C is the minor allele.

[b]C represents the major allele and T is the minor allele.

[c]The *All of Us* Data and Statistics Dissemination Policy prohibits presenting participant counts of fewer than 20.

### Supplementary Figures

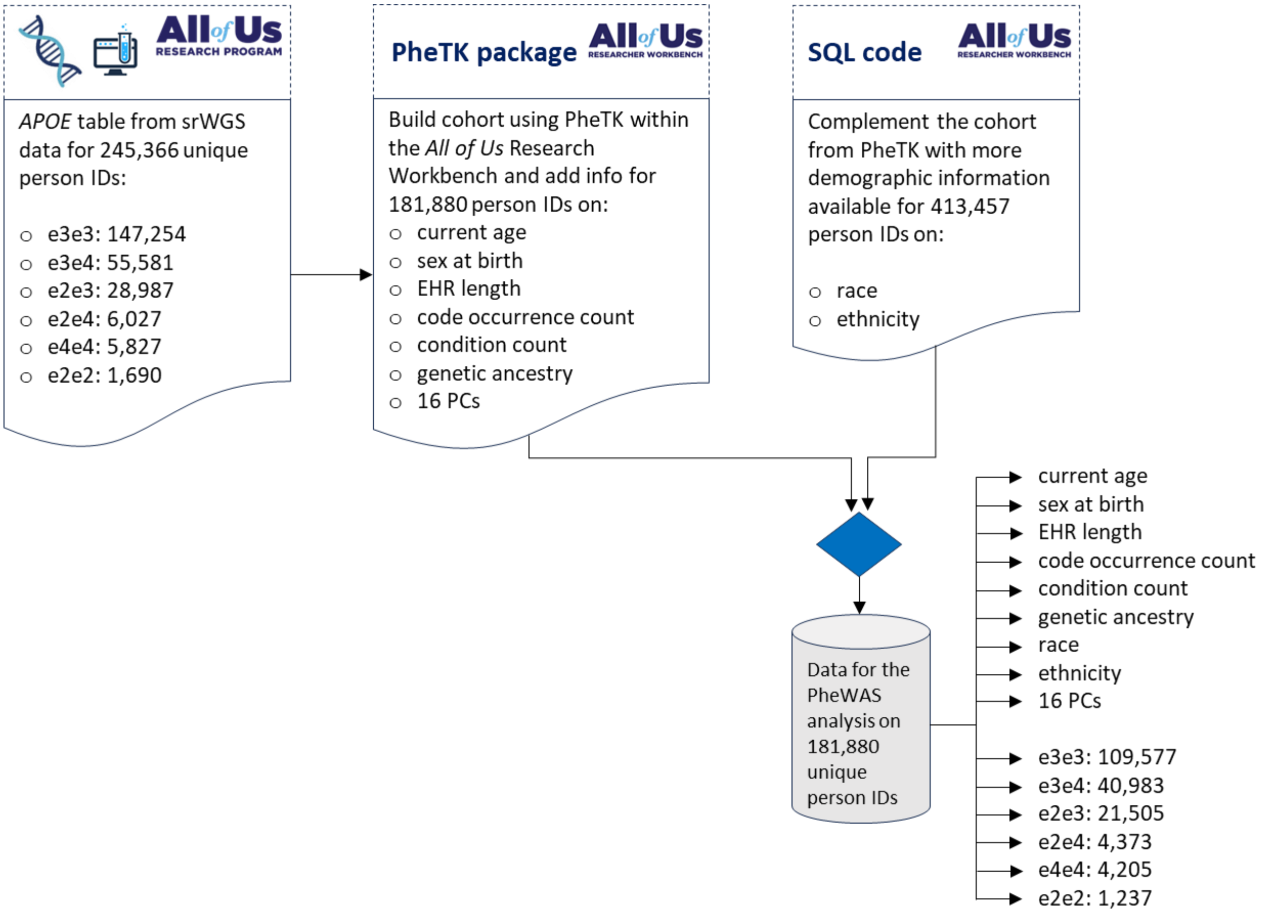


**Figure S1.** Data preparation steps involved in building the study cohort in the *All of Us* Research Program based on the initial 245,366 individuals with the *APOE* genotypic data. The diagram depicts the available phenotypic and genotypic data for the final study population of 181,880 individuals.

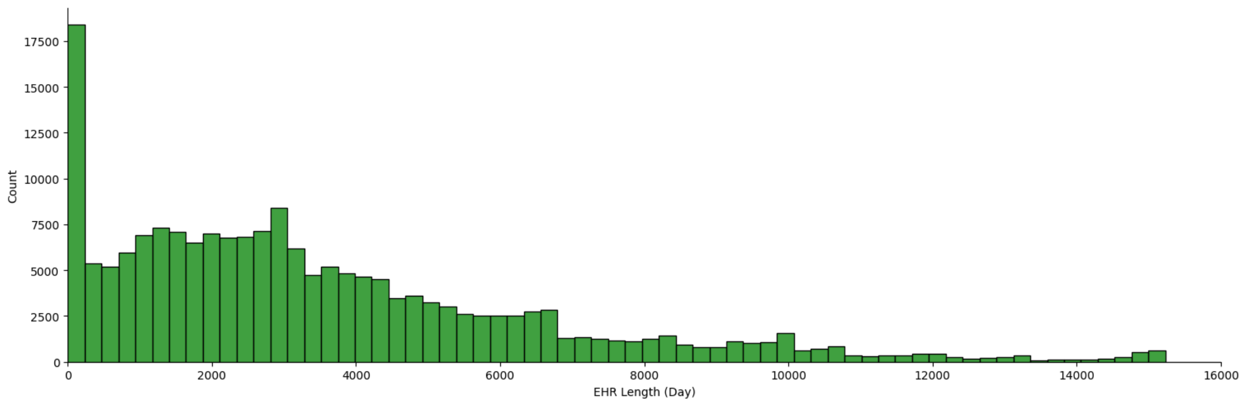


**Figure S2**. Distribution of the EHR length data in days from the overall study population consisted of 181,880 participants. The data was extracted using the PheTK package in the Researcher Workbench. After converting the data to year, the average EHR length was 9.68 years (SD=8.34) in the whole cohort.

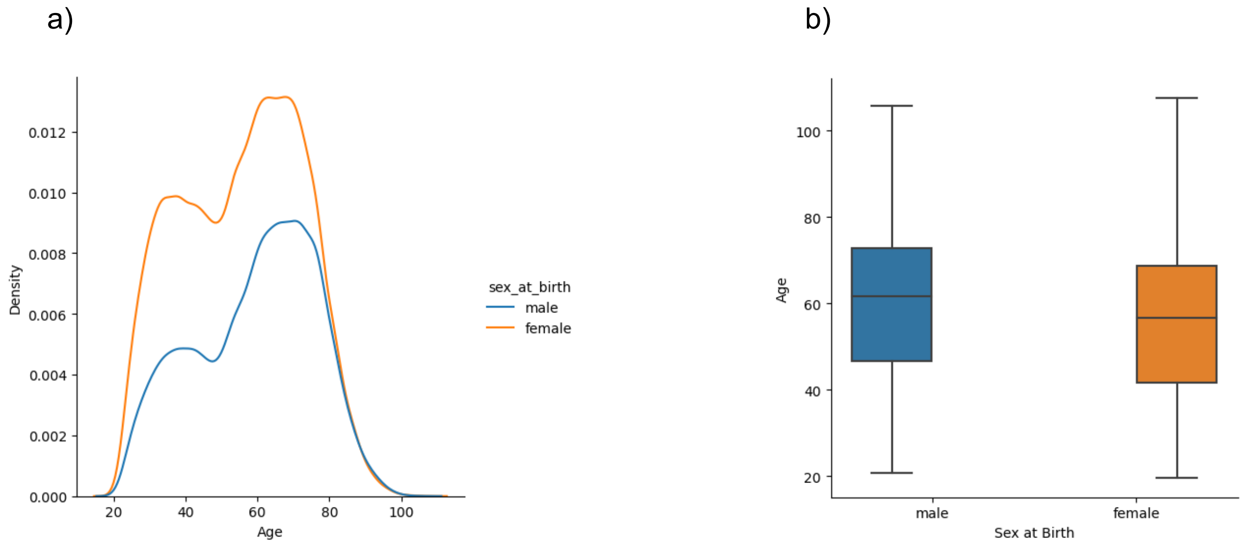


**Figure S3.** Distribution of the participants' age in the whole cohort of 181,880 participants separated based on sex groups assigned at birth for 112,435 females and 69,445 males: a) A density plot displays the distributions b) A box plot depicting the age distributions. The median age of males was 61.68 years, while the median age of females was 56.68 years. Similarly, males on average were older than females, respectively with an average age of 59.35 years (SD=16.79) and average age of 55.46 years (SD=16.84).

a) Females

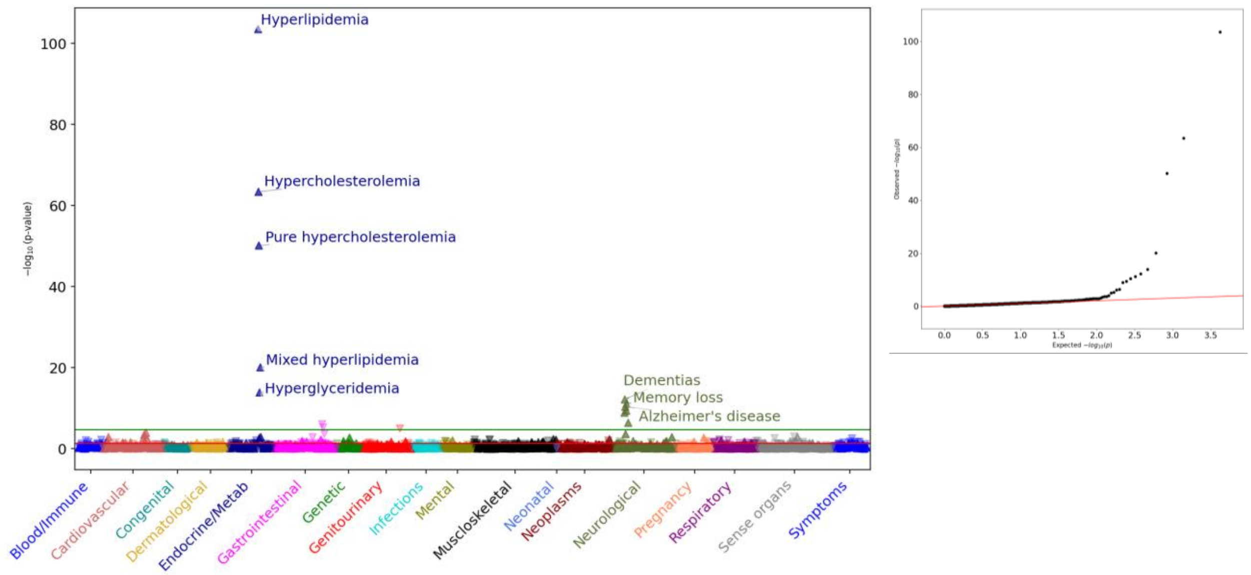


b) Males

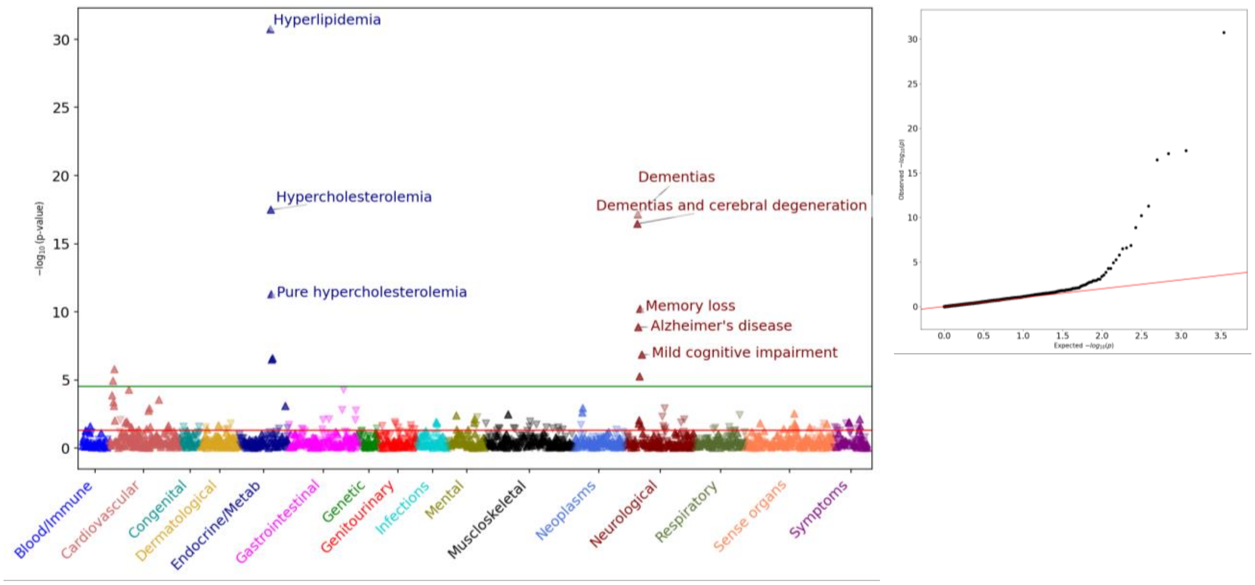


**Figure S4.** PheWAS results by sex assigned at birth, respectively for a) females and b) males, alongside the corresponding QQ plots. The *APOE* variants were arranged from ε2 towards ε4, i.e. ε2/ε2<ε2/ε3<ε3/ε3<ε2/ε4<ε3/ε4<ε4/ε4. Both PheWAS analyses were ordered based on $-\log_{10}$ *P*-values and adjusted for EHR length, age, sex, and PCs 1-16. In the female cohort, 2,096 phecodes with more than 50 cases or controls extracted from 112,435 females were analyzed for association with the *APOE* variants. Likewise, in the male cohort, 1,727 phecodes retrieved from 69,445 males were tested for association. The green horizontal line represents Bonferroni correction level, respectively $\alpha=2.39\times10^{-05}$ (*0.05/2,096 phecodes*) for females and $2.9\times10^{-05}$ for males (*0.05/1,727 phecodes*). The red line reflects the nominal significance ($\alpha=0.05$). Furthermore, the direction of arrows indicates either an increased risk effect of an association or the reverse direction, a decreased risk effect. The x-axis represents the tested phecodes and the broader 18 categories they belong to, and the y-axis shows the *P*-value of estimated effect sizes ($-\log_{10}$ *P*-value). N.B. We note that the y-axis range is not equal in the plots, allowing for better visualization.

a) African/African American (AFR)

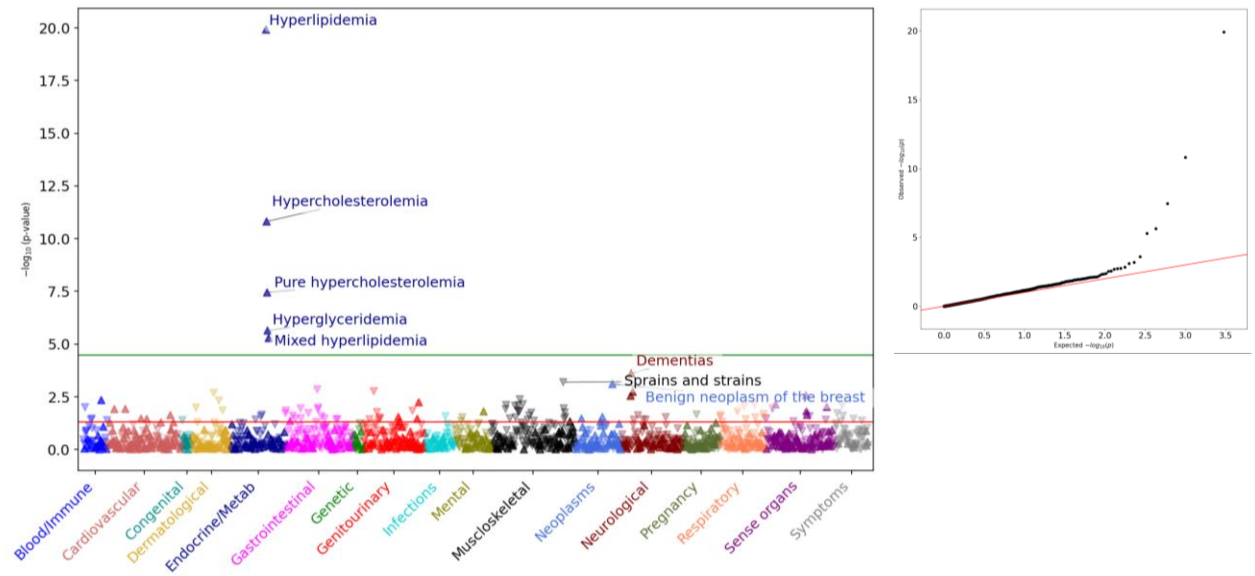


b) American Admixed/Latino (AMR)

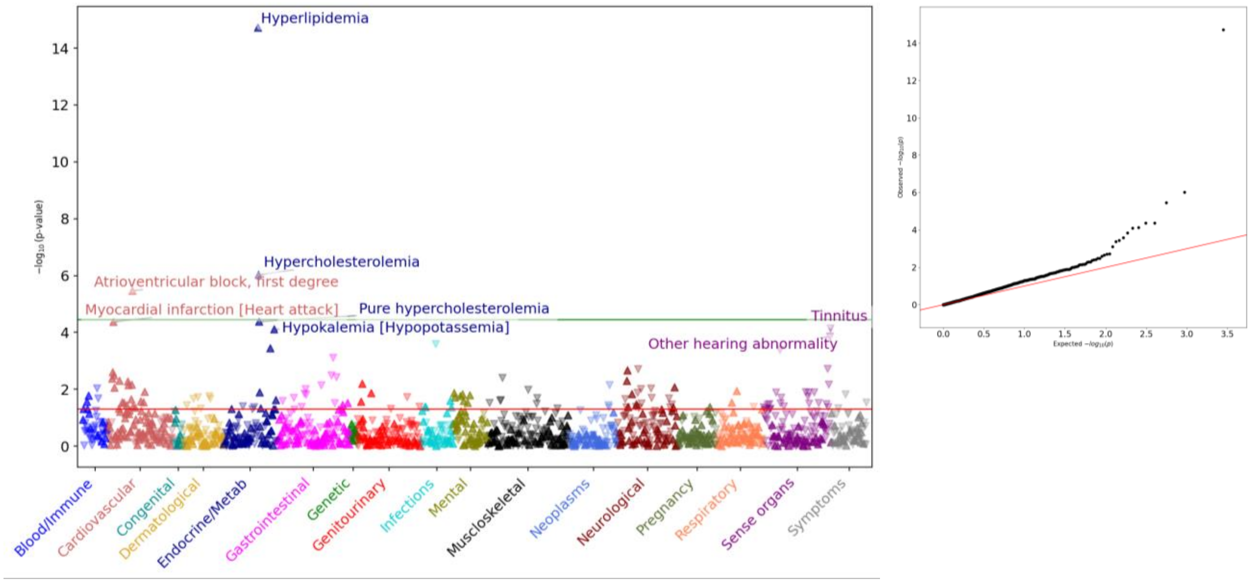

c) East Asian (EAS)

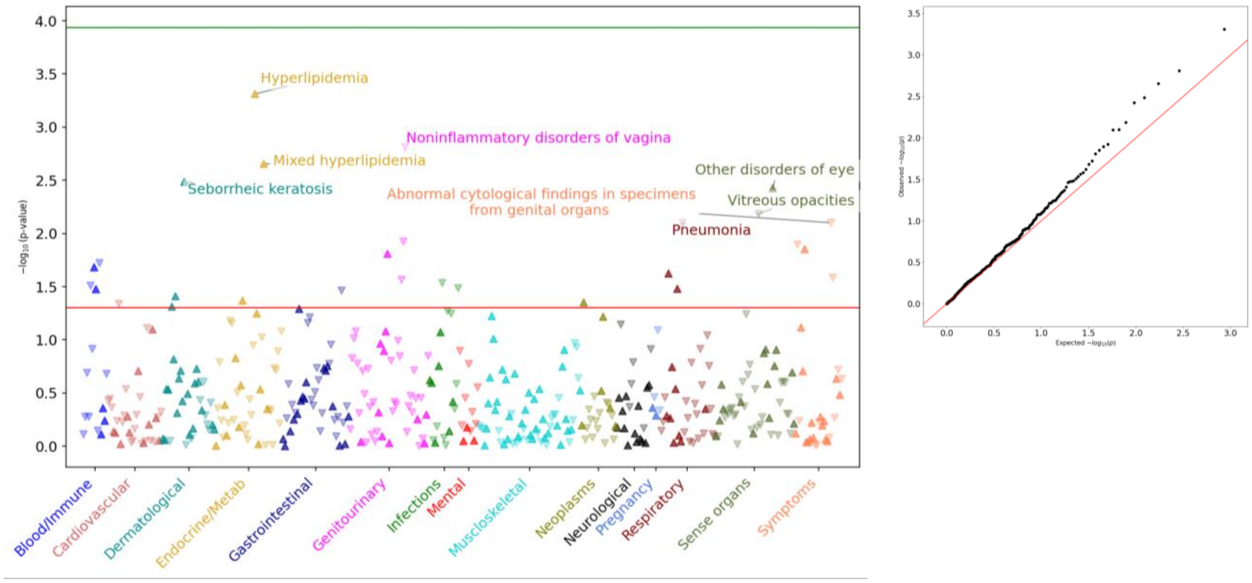


d) European (EUR)

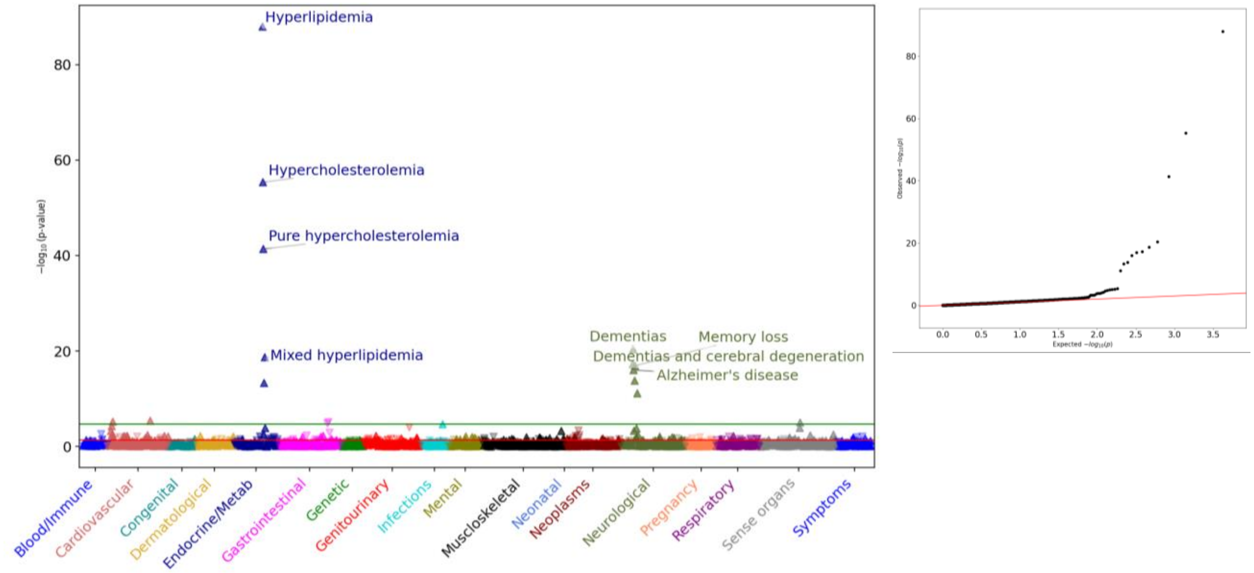

e) Middle Eastern (MID)

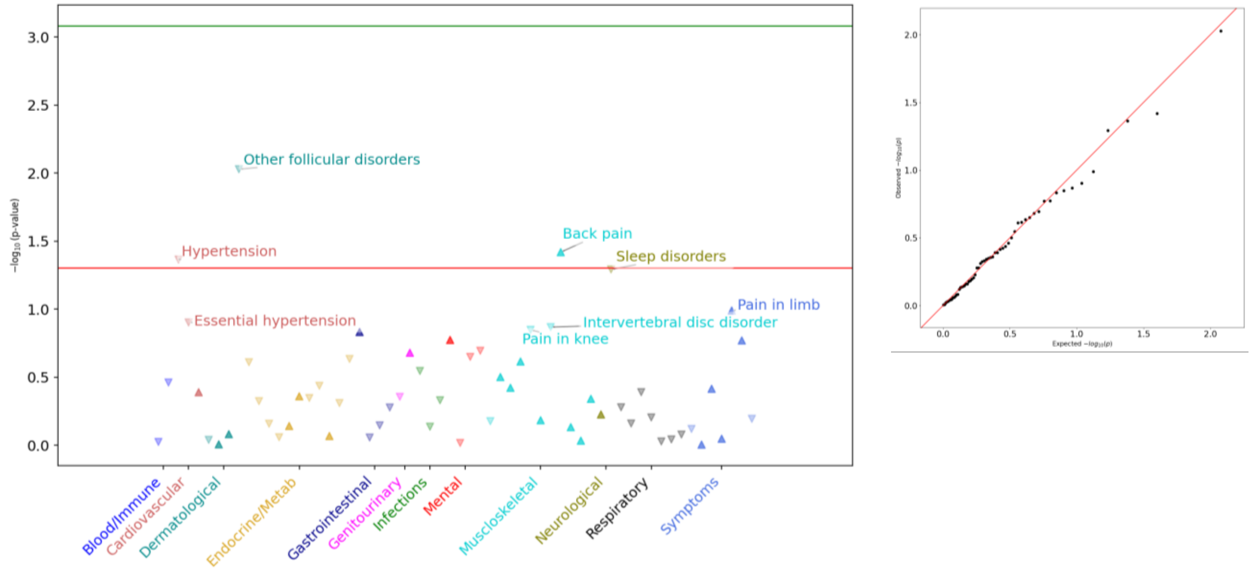


f) South Asian (SAS)

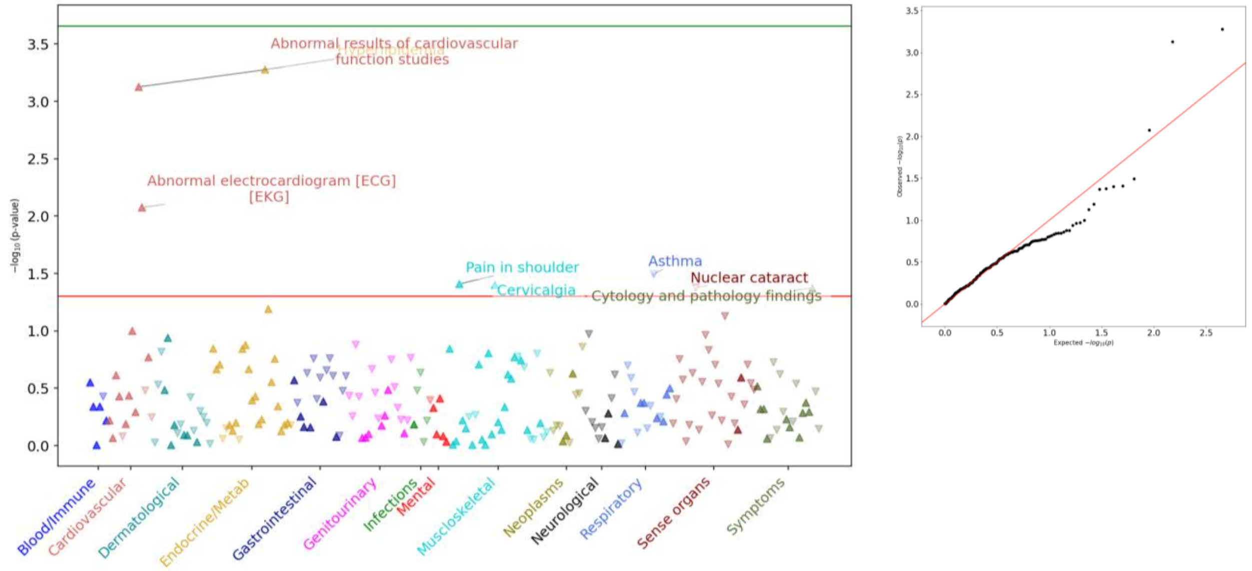


**Figure S5.** PheWAS results and their corresponding QQ plots in ancestral groups presented alphabetically for a) African/African American (AFR) b) American Admixed/Latino (AMR) c) East Asian (EAS) d) European (EUR) e) Middle Eastern (MID), and f) South Asian (SAS). The *APOE* variants were arranged from ε2 towards ε4, i.e. ε2/ε2<ε2/ε3<ε3/ε3<ε2/ε4<ε3/ε4<ε4/ε4. All PheWAS analyses were ordered based on $-\log_{10}$ *P*-values and adjusted for EHR length, age, sex, and PCs 1-16. In the AFR cohort, 1,501 phecodes with more than 50 cases or controls were analyzed for association with the *APOE* variants. Likewise, 1,398, 430, 2,105, 60, and 226 phecodes met the inclusion criteria respectively in the AMR, EAS, EUR, MID, and SAS cohorts and were tested for association. The green horizontal line represents Bonferroni correction level with $\alpha=3.33\times10^{-05}$ (*0.05/1,501 phecodes*) in AFR, $3.58\times10^{-05}$ in AMR, $1.16\times10^{-04}$ in EAS, $2.38\times10^{-05}$ in EUR, $8.33\times10^{-04}$ in MID, and $2.21\times10^{-04}$ in SAS. The red line displays the nominal significance ($\alpha=0.05$). Furthermore, the direction of arrows indicates either an increased risk effect of an association or the reverse direction, a decreased risk effect. The x-axis represents the tested phecodes and the broader 18 categories they belong to, and the y-axis shows the *P*-value of estimated effect sizes (-$\log_{10}$ *P*-value). N.B. We note that the y-axis range is not equal in the plots, allowing for better visualization.

a) *APOE* variants: ε2/ε2<ε2/ε3<ε3/ε3

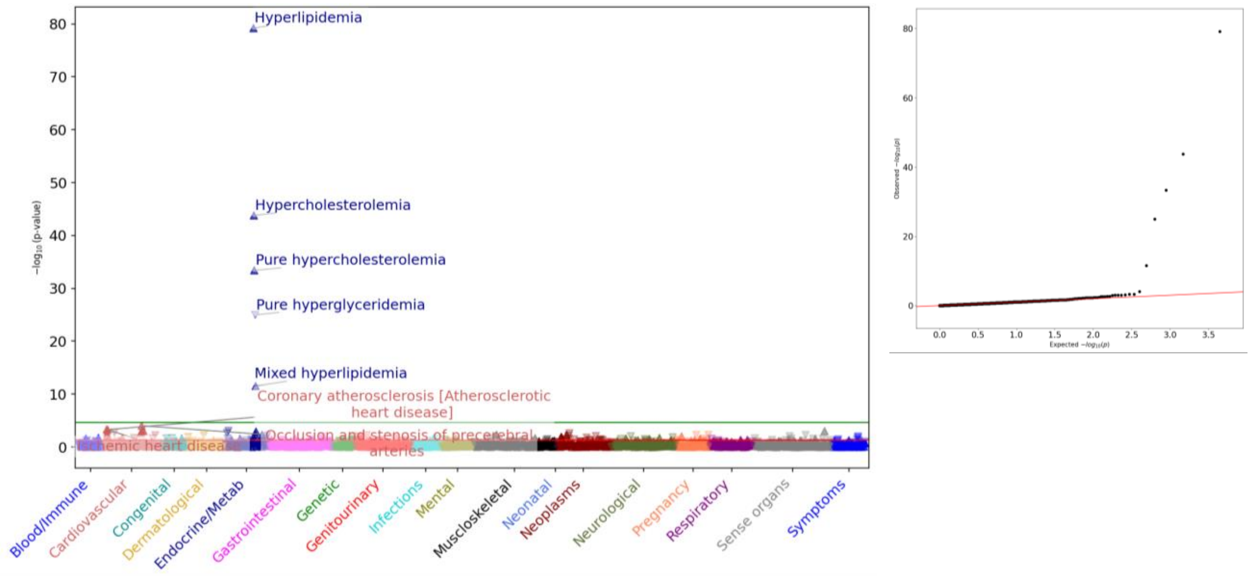


b) *APOE* variants: ε3/ε3<ε3/ε4<ε4/ε4

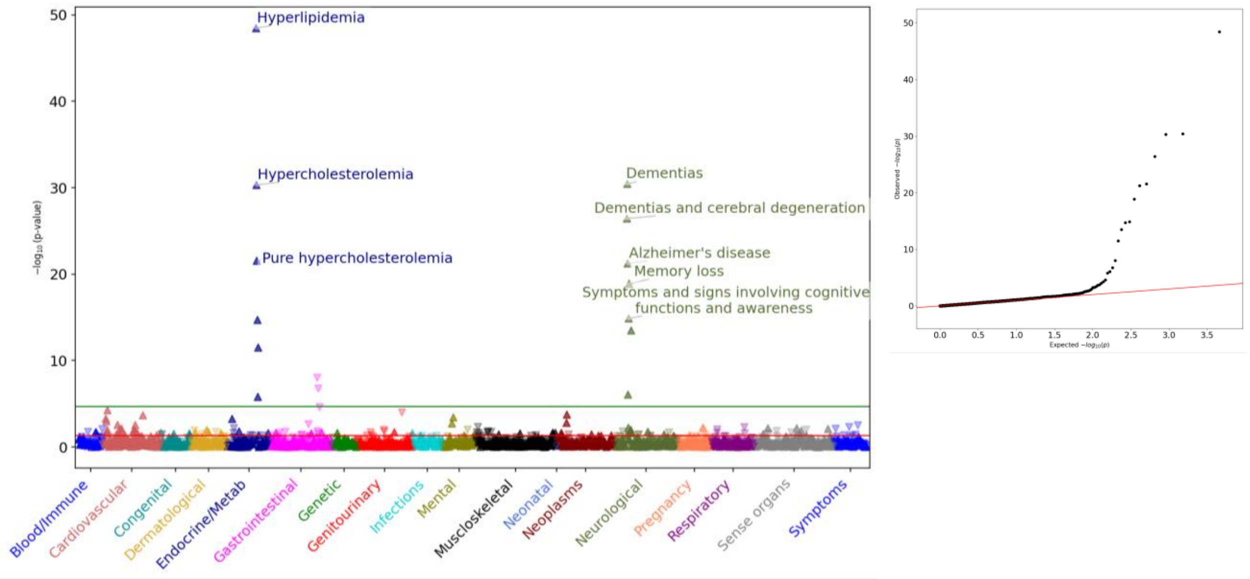


**Figure S6.** PheWAS results by subsets of the *APOE* variants in the whole cohort respectively for a) ε2/ε2<ε2/ε3<ε3/ε3 and b) ε3/ε3<ε3/ε4<ε4/ε4, alongside their corresponding QQ plots. Both PheWAS analyses ordered based on -$\log_{10}$ *P*-values and adjusted for EHR length, age, sex, and PCs 1-16. In the ε2/ε2<ε2/ε3<ε3/ε3 analysis, 2,202 phecodes with more than 50 cases or controls were analyzed for association with the *APOE* variants. Likewise, in the ε3/ε3<ε3/ε4<ε4/ε4 analysis, 2,269 phecodes were tested for association. The green horizontal line represents Bonferroni correction level, respectively $\alpha=2.27\times10^{-05}$ (*0.05/2,202 phecodes*) for the ε2/ε2<ε2/ε3<ε3/ε3 analysis and $2.2\times10^{-05}$ for the ε3/ε3<ε3/ε4<ε4/ε4 analysis (*0.05/2,269 phecodes*). The red line represents the nominal significance ($\alpha$=0.05). Furthermore, the direction of arrows indicates either an increased risk effect of an association or the reverse direction, a decreased risk effect. The x-axis represents the tested phecodes and the broader 18 categories they belong to, and the y-axis shows the *P*-value of estimated effect sizes (-$\log_{10}$ *P*-value). N.B. We note that the y-axis range is not equal in the plots, allowing for better visualization.

a) *APOE* variants: ε3/ε3<ε3/ε4

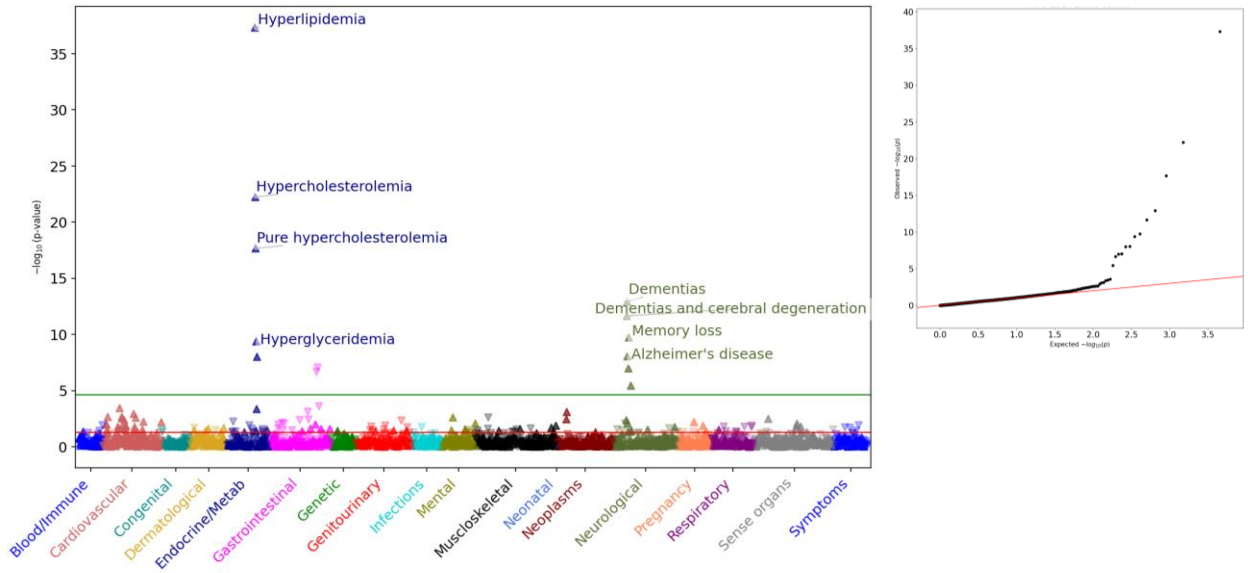


b) *APOE* variants: ε3/ε3<ε4/ε4

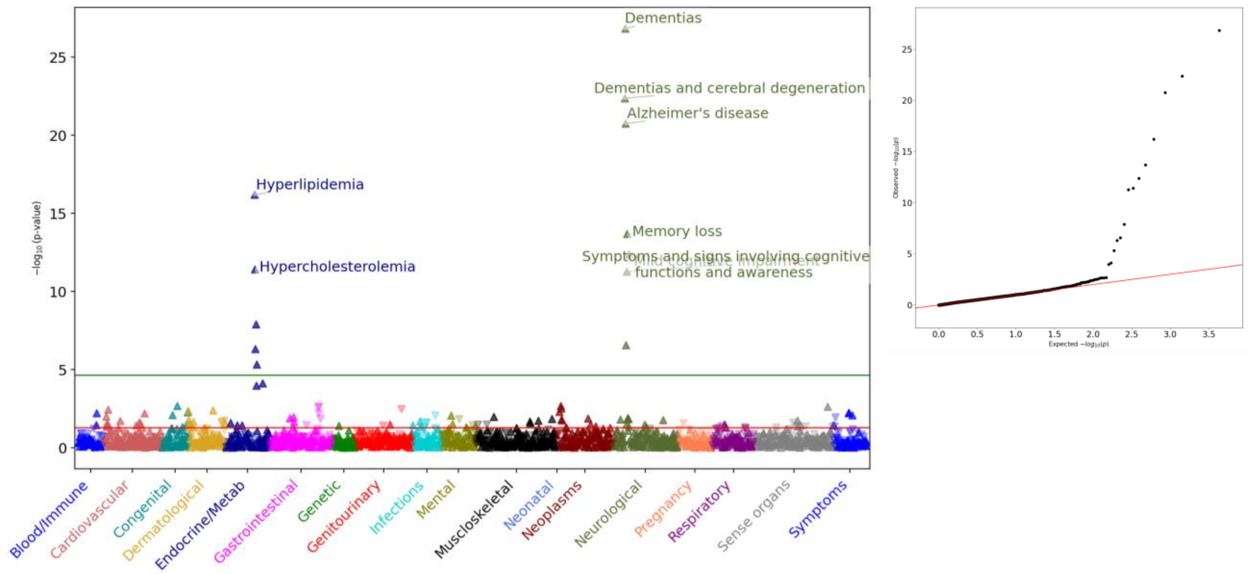


**Figure S7.** PheWAS results by subsets of the *APOE* variants in the whole cohort respectively for a) ε3/ε3<ε3/ε4 and b) ε3/ε3<ε4/ε4, alongside their corresponding QQ plots. Both PheWAS analyses ordered based on $-\log_{10}$ *P*-values and adjusted for EHR length, age, sex, and PCs 1-16. In the first analysis, 2,251 phecodes with more than 50 cases or controls were analyzed for association with the *APOE* variants. Likewise, in the ε3/ε3<ε4/ε4 analysis, 2,141 phecodes were tested for association. The green horizontal line represents Bonferroni correction level, respectively $\alpha=2.22\times10^{-05}$ (*0.05/2,251 phecodes*) for the ε3/ε3<ε3/ε4 analysis and $2.34\times10^{-05}$ for the second analysis (*0.05/2,141 phecodes*). The red line represents the nominal significance ($\alpha=0.05$). Furthermore, the direction of arrows indicates either an increased risk effect of an association or the reverse direction, a decreased risk effect. The x-axis represents the tested phecodes and the broader 18 categories they belong to, and the y-axis shows the *P*-value of estimated effect sizes ($-\log_{10}$ *P*-value). N.B. We note that the y-axis range is not equal in the plots, allowing for better visualization.

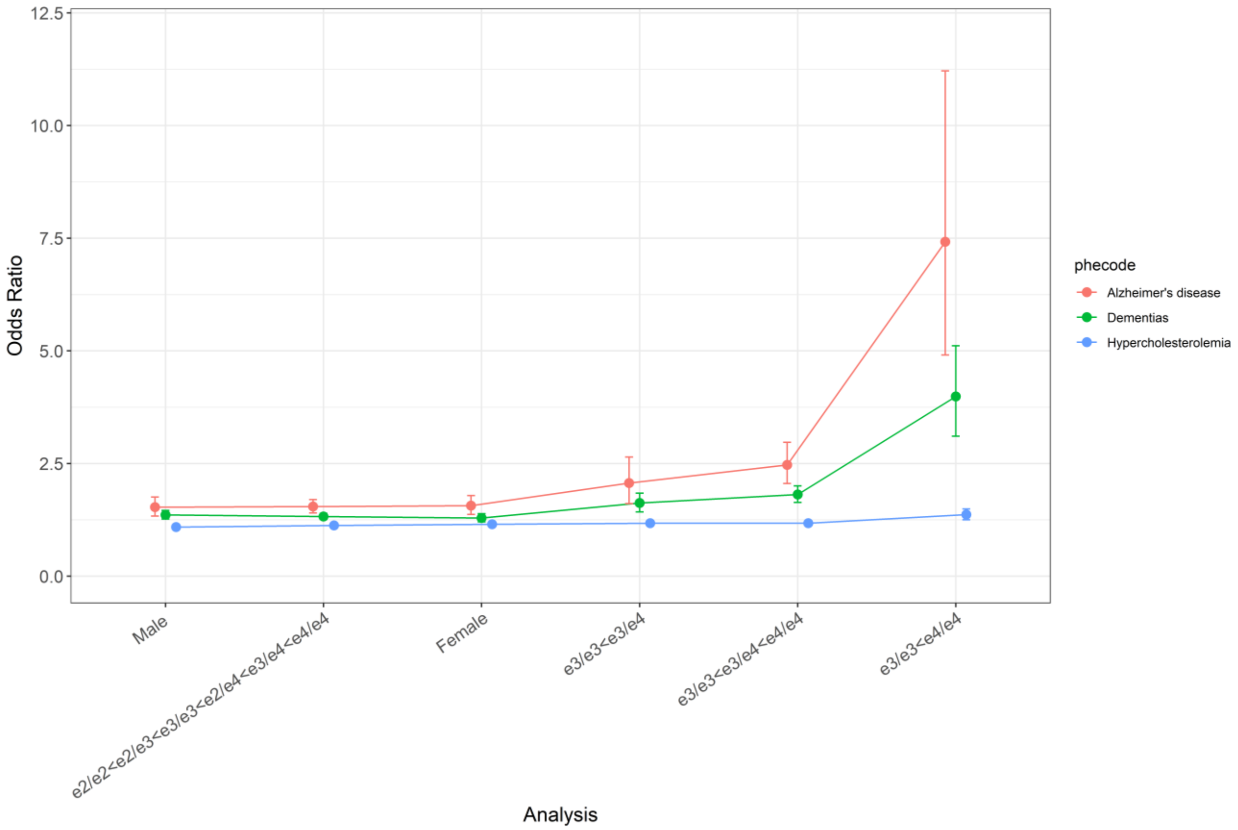


**Figure S8.** Odds ratios (ORs) with confidence intervals are presented for three phecodes that consistently appeared in the top associations with *APOE* variants across distinct analyses in the present study. The ε3/ε3<ε4/ε4 analysis yielded the highest ORs for all three phecodes across the six analyses, and in the meantime, AD possessed higher ORs than the other phecodes in all comparisons.
